# Ethnicity and anthropometric deficits in children: a cross-sectional analysis of national survey data from 18 countries in sub-Saharan Africa

**DOI:** 10.1101/2023.10.10.23296801

**Authors:** Lucy S. Tusting, Harry S. Gibson, Swapnil Mishra, Steven W. Lindsay, Daniel J. Weiss, Seth Flaxman, Samir Bhatt

**Affiliations:** Department of Disease Control, London School of Hygiene & Tropical Medicine, London, UK; Big Data Institute, Nuffield Department of Medicine, University of Oxford, Oxford, UK; Saw Swee Hock School of Public Health, National University of Singapore, Singapore; Department of Biosciences, Durham University, Durham, UK; Telethon Kids Institute, Perth, Australia; Curtin University, Perth, Australia; Department of Computer Science, University of Oxford, UK; Department of Infectious Disease Epidemiology, Imperial College London, London, UK; Section of Epidemiology, Department of Public Health, University of Copenhagen, Copenhagen, Denmark

## Abstract

**Background:** Anthropometric deficits persist in sub-Saharan Africa (SSA) despite sustained improvements in nutrition, disease burden and living conditions. The UN Sustainable Development Goals advocate for disaggregation of health indicators by ethnic group. However, few studies have assessed how ethnicity is associated with anthropometric deficits across SSA.

**Methods:** Data were extracted from 37 georeferenced Demographic and Health Surveys carried out during 2006-2019 across SSA that recorded anthropometric data for children aged <5 years. In a cross-sectional analysis, the odds of stunting (low height-for-age), wasting (low weight-for-height) and underweight (low weight-for-age) were modelled in relation to ethnic group using a generalised linear hierarchical mixed-effects model, controlling for survey design and environmental, socioeconomic, and clinical variables.

**Findings:** The study population comprised 138,312 children spanning 45 ethnic groups across 18 countries. In pairwise comparisons between ethnic groups, height-for-age Z scores differed by at least 0.5 standard deviations in 56% of comparisons, weight-for-height Z scores in 39% of comparisons and weight-for-age Z scores in 34% of comparisons. Compared to a reference group of Fula children (the largest ethnic group), ethnic group membership was associated with both increases and decreases in growth faltering, ranging from a 69% reduction to a 32% increase in odds of stunting (Igbo: adjusted odds ratio (aOR) 0.31, 95% confidence intervals (CI) 0.27-0.35, p<0.0001; Hausa: aOR 1.32, 95% CI 1.21-1.44, p<0.0001); a 13% to 87% reduction in odds of wasting (Mandinka: aOR 0.87, 95% CI 0.76-0.99, p=0.034; Bamileke: aOR 0.13, 95% CI 0.05-0.32, p<0.0001) and an 85% reduction to 13% increase in odds of underweight (Bamileke: aOR 0.15, 95% CI 0.08-0.29, p<0.0001; Hausa: aOR 1.13, 95% CI 1.03-1.24, p=0.010).

**Interpretation:** Major ethnic disparities in stunting, wasting and underweight were observed across 18 countries in SSA. Understanding and accounting for these differences is essential to support progress monitoring and targeting of nutrition interventions in children.

**Funding:** UK Medical Research Council, Novo Nordisk Foundation

**Research in context:** *Evidence before this study:* We searched PubMed with no date restrictions for studies published in English, using the following search terms: (“child*”, “five” OR “infant”) AND (“child growth”, “stunting”, “stunted”, “growth failure”, “growth faltering”, “height” OR “anthropometric”) AND (“ethnic*”). We identified 288 studies (196 from the database search and 92 from reference lists). Of 93 studies full text studies screened, 37 were relevant. Two multi-country studies measured the association between ethnicity and growth outcomes. An analysis of 13 national surveys from Latin America during 2006-2020 found a 97% higher prevalence of stunting among indigenous than European or mixed ancestry participants. In a 2014 systematic review, 20% of height means in 55 countries or ethnic groups differed by ≥0.5 standard deviations (SD) from the WHO Multicentre Growth Reference Study mean, suggesting some differences. A further 35 local studies measured ethnicity as a potential risk factor for child growth outcomes in Australia, Brazil, China, Guatemala, Hawaii, India, Iran, Lithuania, Malaysia, Nepal, Peru, South Africa, Thailand, Trinidad and Tobago, the UK and the USA, with a range of associations observed. We identified additional multi-country, population-based cohorts designed to support the development of international growth standards, but these did not specifically measure inequalities between ethnic groups.

*Added value of this study:* To our knowledge, this is the first systematic, multi-country analysis of ethnicity and anthropometric deficits in sub-Saharan Africa. By analysing data for 138,312 children spanning 45 ethnic groups in 18 countries, measured in 37 Demographic and Health Surveys, we found ethnicity to be a primary risk factor for anthropometric deficits after adjusting for socioeconomic, environmental and child-level characteristics. The strength of this association exceeded that for other factors known to affect children’s growth, such as household wealth, history of diarrhoea and access to improved water and sanitation. Anthropometric z-scores differed by ≥0.5 SD (a clinically relevant threshold) in 34%-56% of pairwise comparisons between ethnic groups.

*Implications of all the available evidence:* Child growth faltering persists as a major cause of morbidity and mortality in sub-Saharan Africa^1^ but our study shows that this burden is unequally distributed among ethnic groups. Research is needed to understand these differences, in order to target interventions and effectively track progress towards Sustainable Development Goal 2.

## Introduction

Childhood anthropometric deficits persist globally despite reductions since 2000. The Global Nutrition Targets, incorporated into the 2030 Sustainable Development Goals (SDGs), aim to reduce by 2025 the number of stunted children aged <5 years by 40% and the prevalence of early childhood wasting to below 5%.^2^ However, fewer than half of low- and middle-income countries (LMICs) are expected to reach this goal.^3^ In sub-Saharan Africa (SSA), poor growth outcomes present a major public health challenge^1^ with an estimated 37% of children aged <5 years stunted (insufficient height-for-age), 9% wasted (insufficient weight-for-height) and 20% underweight (insufficient weight-for-age) in 2015.^3^ These deficits exacerbate other primary causes of childhood morbidity and mortality in SSA, ultimately accounting for an estimated 23% of deaths of children aged <5 years in this region in 2015.^4^

Quantitative measurements of trends and distribution of disease burden are necessary to measure progress towards the SDGs and improve intervention targeting. For indicators of children’s growth including stunting, wasting and underweight, high-resolution estimates of national^4^ and sub-national^3^ disease burden have been produced. However, the SDGs also advocate for examination of progress towards individual goals disaggregated by socioeconomic and demographic variables. Ethnicity, which defines a large population group with a shared culture, language and heritage, is a key factor associated with health risks and outcomes.^5^ Child growth outcomes have been investigated relative to ethnicity in two multi-country studies,^6,7^ and compared between populations in pooled international analyses.^8–10^ Growth outcomes have also been measured in multi-country, population-based cohorts designed to support the development of international growth standards (including the WHO MGRS study^11^ and the INTERGROWTH-21^st^ study^12^); however, these studies did not specifically investigate inequalities between ethnic groups. To our knowledge, no studies have systematically measured the association between ethnicity and anthropometric deficits across SSA. Here we investigated whether there are significant differences in anthropometric deficits among children aged <5 years belonging to different ethnic groups across SSA using nationally representative survey data, adjusting for environmental, socioeconomic and clinical covariables.

## Materials and Methods

### Data sources

Data were extracted from the Demographic and Health Survey (DHS) program. DHS surveys are conducted every 3-5 years using a two-stage random cluster sampling strategy to collect nationally representative health and sociodemographic data.^13^ We included any georeferenced surveys conducted in SSA that were published by February 2021 and collected anthropometric data and all prespecified covariables (Figure 1).

**Figure 1.**
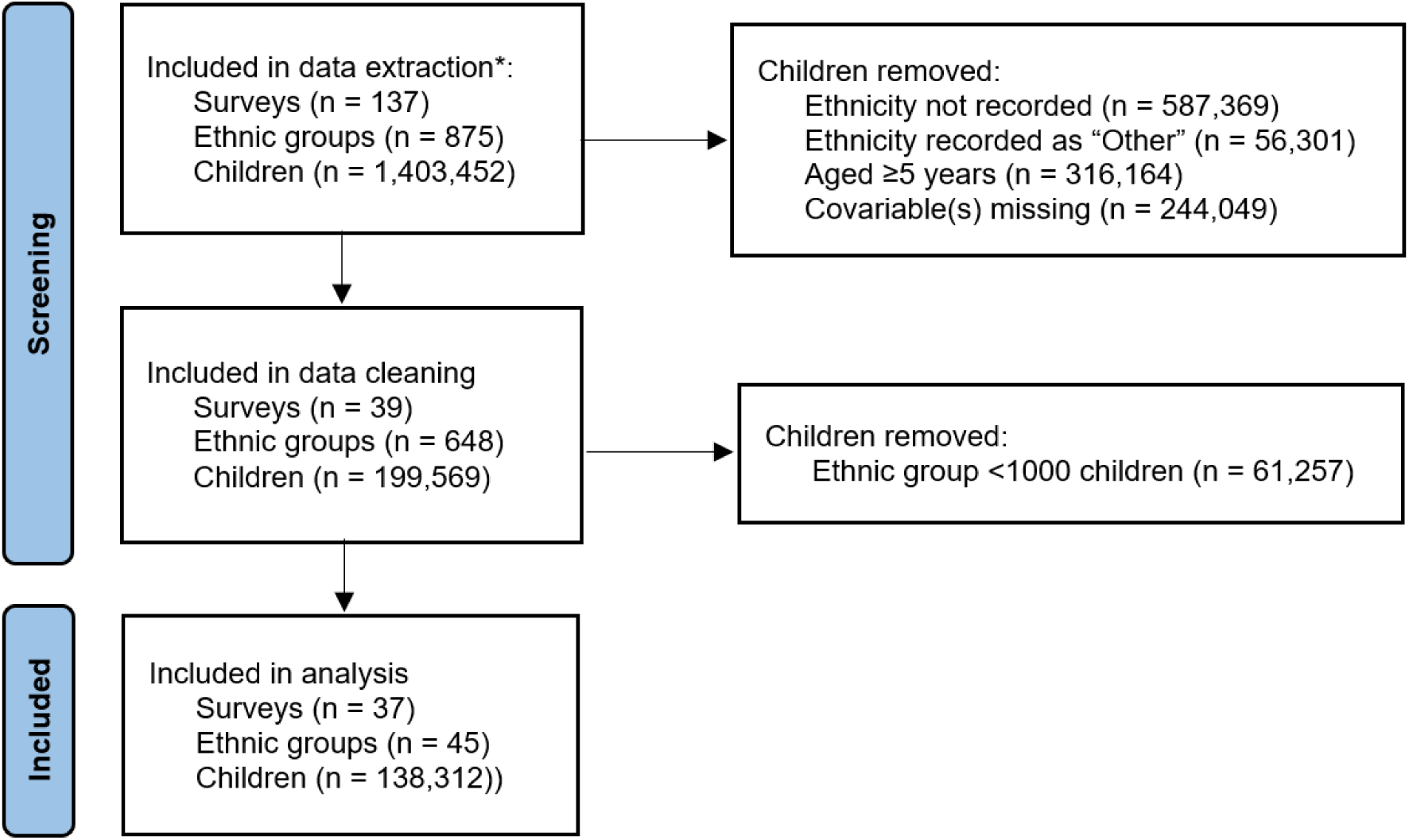
Study flow. All georeferenced surveys from sub-Saharan Africa that were published by February 2021 and collected anthropometric data and all prespecified covariables were included in the data extraction.*

#### Growth outcomes

Anthropometric data were extracted for children aged <5 years. Stunting was defined as a height-for-age z-score (HAZ) greater than two standard deviations (SD) below the WHO 2006 reference median, wasting as a weight-for-height z-score (WHZ) lower than -2 SD, and underweight as a weight-for-age z-score (WAZ) lower than -2 SD. WHO 2006 criteria for cleaning z-scores were applied, where children were excluded if they had HAZ scores below -6 SD or above +6 SD, WAZ scores below -6 SD or above +5 SD, or WHZ scores below -5 SD or above +5 SD.^14^ Like WHO, we used 0.5 SD as a benchmark for clinically significant differences.^11^

#### Ethnicity

Within survey households, the mother or primary caregiver of each child provided self-reported ethnicity. The named ethnicity was assumed to apply to each child. Ethnic groups with fewer than 1,000 children were excluded to prevent spurious relationships from low sample sizes and to limit the overall number of ethnic groups. The largest ethnic group (Fula) was used as the reference group to prevent a rank deficient design matrix.

#### Child and household characteristics

The following individual-level covariables were extracted for each child: age, gender, reported use of an insecticide-treated net (ITN) the previous night, receipt of the third diphtheria-pertussis-tetanus (DPT-3) vaccination, receipt of the first measles vaccination (measles-1), any reported episode of diarrhoea in the previous two weeks, whether or not the child was exclusively breastfed (for children aged <1 year), and whether height was measured standing or lying. Child-level covariables were linked with the following household-level covariables: improved or unimproved drinking water source and sanitation facility categorised using WHO Joint Monitoring Programme (WHO-JMP) criteria ^15^ (i.e. for drinking water, whether or not the source has adequate protection from outside contamination – improved sources include piped water, protected wells and rainwater; for sanitation facility, whether or not the facility adequately separates human excreta from human contact), education level of the household head, urban or rural residence, and finished versus unfinished or natural floor. Since household wealth scores were missing for some surveys, a household wealth index was re-calculated across all surveys using linear principal component analysis using methods described elsewhere.^16^ The following assets were included: bicycle, car, cart, electricity, landline telephone, mobile telephone, motorboat, radio, refrigerator, scooter, television and watch. However, for each survey we excluded assets with more than 10% missing values and a population frequency <5% or >95%.

#### Climate and environmental variables

For each survey cluster the following data were extracted: synoptic mean monthly daytime land surface temperature (LST) from 2000 to 2016 (i.e. the average temperature for each month derived from a multiyear time series);^17^ synoptic mean monthly enhanced vegetation index (EVI) from 2000 to 2020, which is correlated with vegetation density;^17^ synoptic total monthly rainfall from 2000 to 2020^18^ and accessibility to large cities in 2015.^19^

#### Association between stunting and ethnicity

The association between ethnicity and six growth outcomes (stunting, wasting, underweight, HAZ score, WHZ score and WAZ score) was modelled using a generalised linear hierarchical mixed effects model. The model included a fixed effects design matrix (X) for age and gender of the child, ITN use, DPT-3 vaccination, measles-1 vaccination, reported diarrhoea, height measurement position, drinking water source, sanitation facility, secondary education of the household head, urban or rural survey cluster, household floor material, household wealth, monthly mean LST, monthly mean EVI, mean total monthly rainfall and accessibility to large cities. Breastfeeding status was not included in the model due to high missing data. Except for age, continuous fixed effects were standardised to make them directly interpretable with binary fixed effects. The linear predictor of z-scores and binary growth outcomes for a given child i was modelled as:

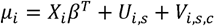

Where the terms *U_i,s_* are survey random effects, *V_i,s,c_* are survey-cluster random effects and *X_i_* is the design matrix of covariables. Conceptually, the covariables account for the effect of wealth, education and other factors, the survey effects provide an adjustment factor based on the survey and the survey-cluster effects provide a further adjustment to a specific cluster within a given survey. A normal likelihood was used for fitting z-scores and a binomial likelihood for binary growth outcomes. Fitting was performed using glmmTMB via restricted maximum likelihood.^20^ Standard errors and confidence intervals were estimated using the Wald method.

#### Model performance

The predictive performance of the model was assessed using out-of-sample validation, by random partitioning of 10,000 children into a validation dataset and the remaining observations into a training dataset. This scheme was repeated 10 times and scores averaged. We evaluated the model at both the child and aggregated ethnicity level. Mean squared error and the correlation coefficient were used to evaluate model performance for z-scores. Area under a Receiver Operating Characteristic curve was used to assess specificity of predictions for binary growth outcomes at child level.

#### Role of the funding source

The study sponsors had no role in the study design; in the collection, analysis, and interpretation of data; in the writing of the report; or in the decision to submit the paper for publication.

## Results

### Study population

Descriptive statistics are shown in S1 Table and S2 Table. Data were extracted for 137 AIDS, DHS, MIS and SPE surveys in Asia, Latin America and SSA, of which 37 DHS surveys that measured all pre-specified variables were analysed (Figure 1). The included surveys were conducted during 2006-2019 in 18 countries, all of which were in SSA. A total of 138,312 children had complete data and belonged to an ethnic group with at least 1,000 observations. These children were resident in 92,588 households spanning 45 ethnic groups, the largest being Fula (n=17,558 children, 12.7%). Median cluster size was 10 households (interquartile range 6, 14). The mean age of children was 2.2 years (95% confidence intervals (CI) 2.2, 2.2) and 69,478 (50.2%) children were male.

### Association between stunting and ethnicity

Overall, 46,135 (33.4%) of 138,312 children were stunted and 81% had a height-for-age lower than the WHO median. Stunting prevalence ranged from 17.8% (1,026 of 5,752) in Igbo children to 52.3% (6,555 of 12,540) in Hausa children (S3 Table, S4 Table). Greater variation in HAZ scores was found between, than within, ethnic groups (p<0.0001) (Figure 2, Figure 3a). HAZ scores differed by at least 0.5 SD in 56% of significant pairwise comparisons between ethnic groups. The largest difference between any two ethnic groups was 1.30 SD (Figure 3b). Compared to Fula children, the adjusted difference in HAZ score associated with ethnicity ranged from 0.84 SD higher (95% CI 0.75-0.93, p<0.0001) in Igbo children to 0.30 SD lower (95% CI -0.59, -0.00, p=0.047) in Bemba children. Compared to Fula children, the adjusted difference in odds of stunting associated with ethnicity ranged from 69% lower odds in Igbo children (adjusted Odds Ratio (OR) 0.31, 95% CI 0.27-0.35, p<0.0001) to 32% higher odds in Hausa children (aOR 1.32, 95% CI 1.21-1.44, p<0.0001) (Figure 4, S5 Table). As a comparison, the association between stunting and other covariables ranged from 31% lower odds of stunting in wealthier compared to less wealthy households (aOR 0.69, 95% CI 0.67-0.72; p<0.0001) to 30% higher odds of stunting in males compared to females (aOR 1.30, 95% CI 1.27-1.33, p<0.0001).

**Figure 2.**
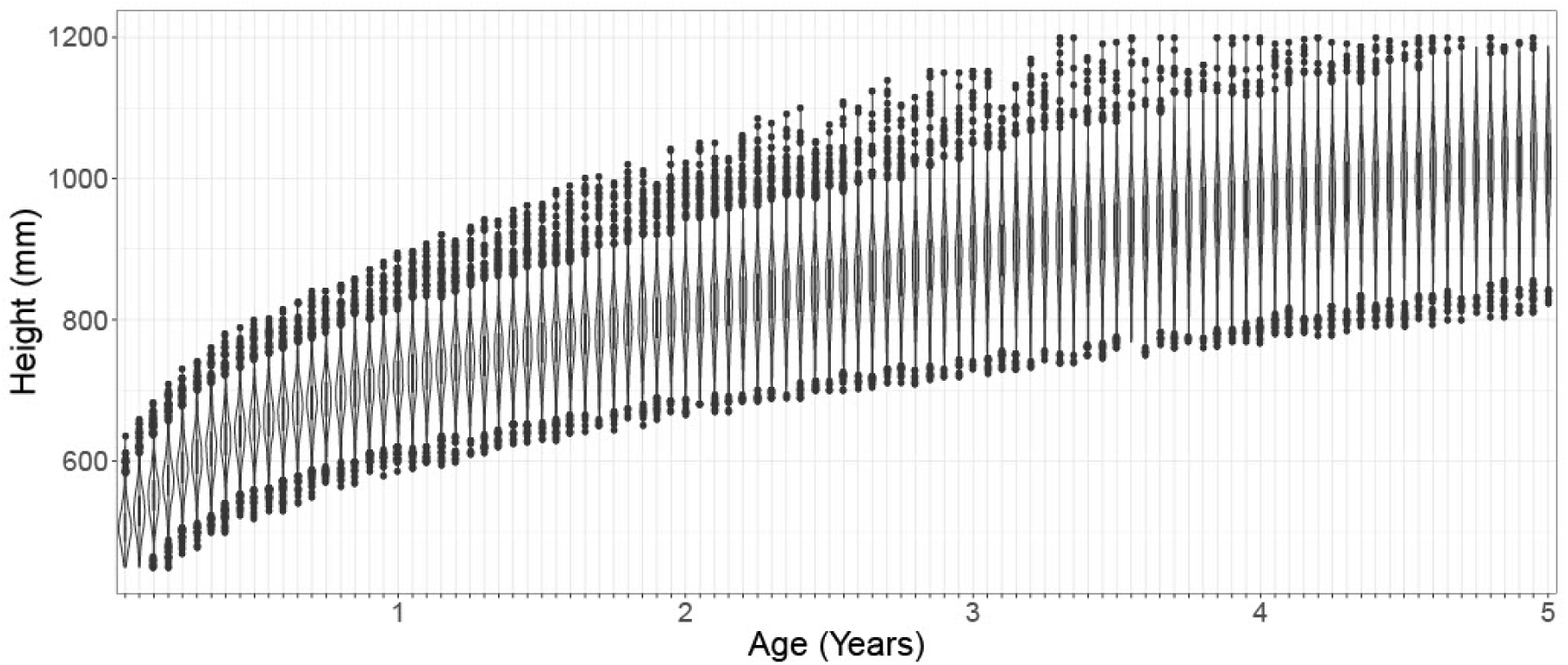
Variation in height-for-age by ethnicity. Points represent median of absolute height-for-age for each ethnic group.

**Figure 3.**
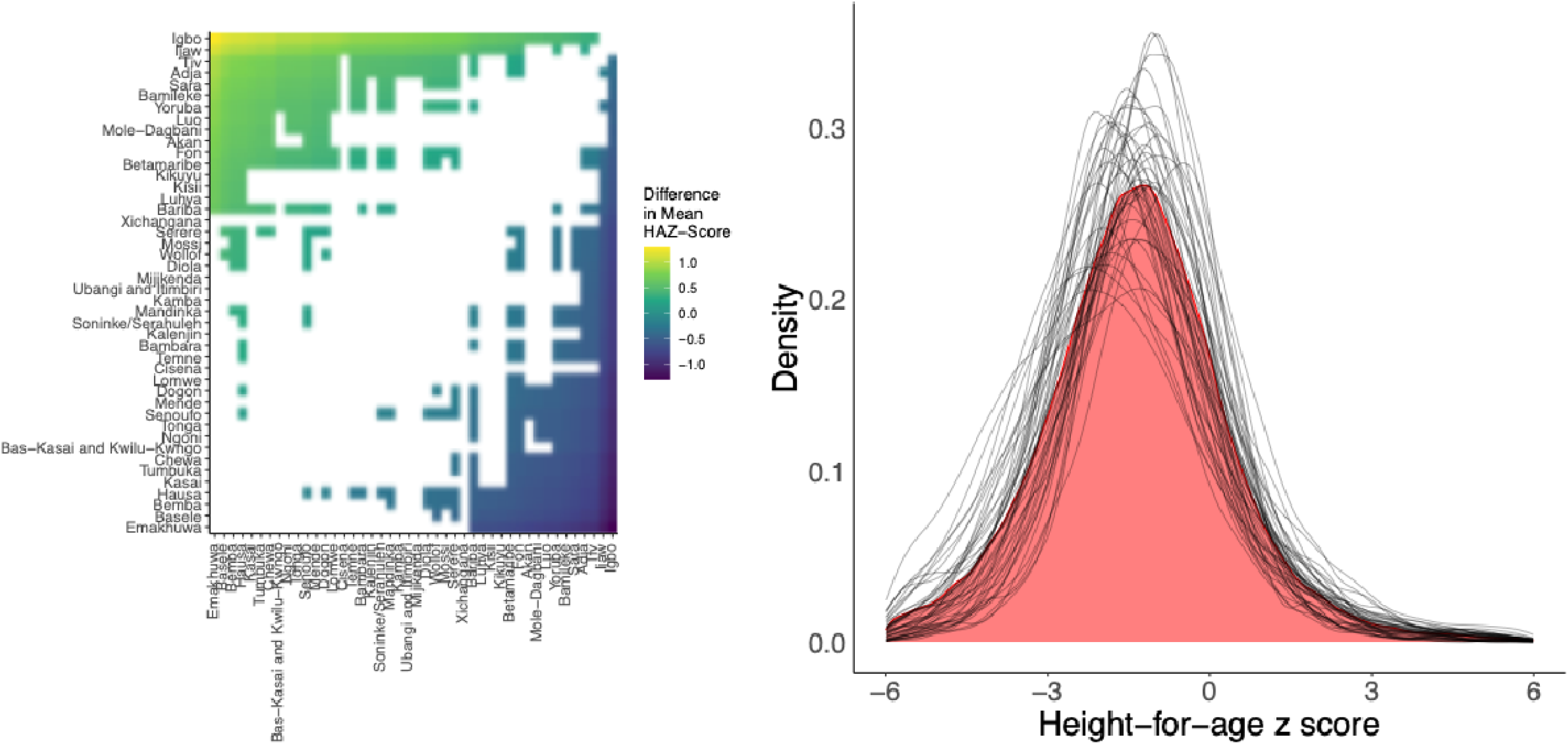
Growth variation relative to ethnic group among 138,312 children aged <5 years in 18 countries in sub-Saharan Africa surveyed between 2006 to 2019. Right: Density plot shows distribution of height-for-age Z (HAZ) scores by ethnic group, illustrating variation in growth patterns between ethnicities. Left: significant (T-test) pair-wise comparisons of mean HAZ scores between ethnic groups.

**Figure 4.**
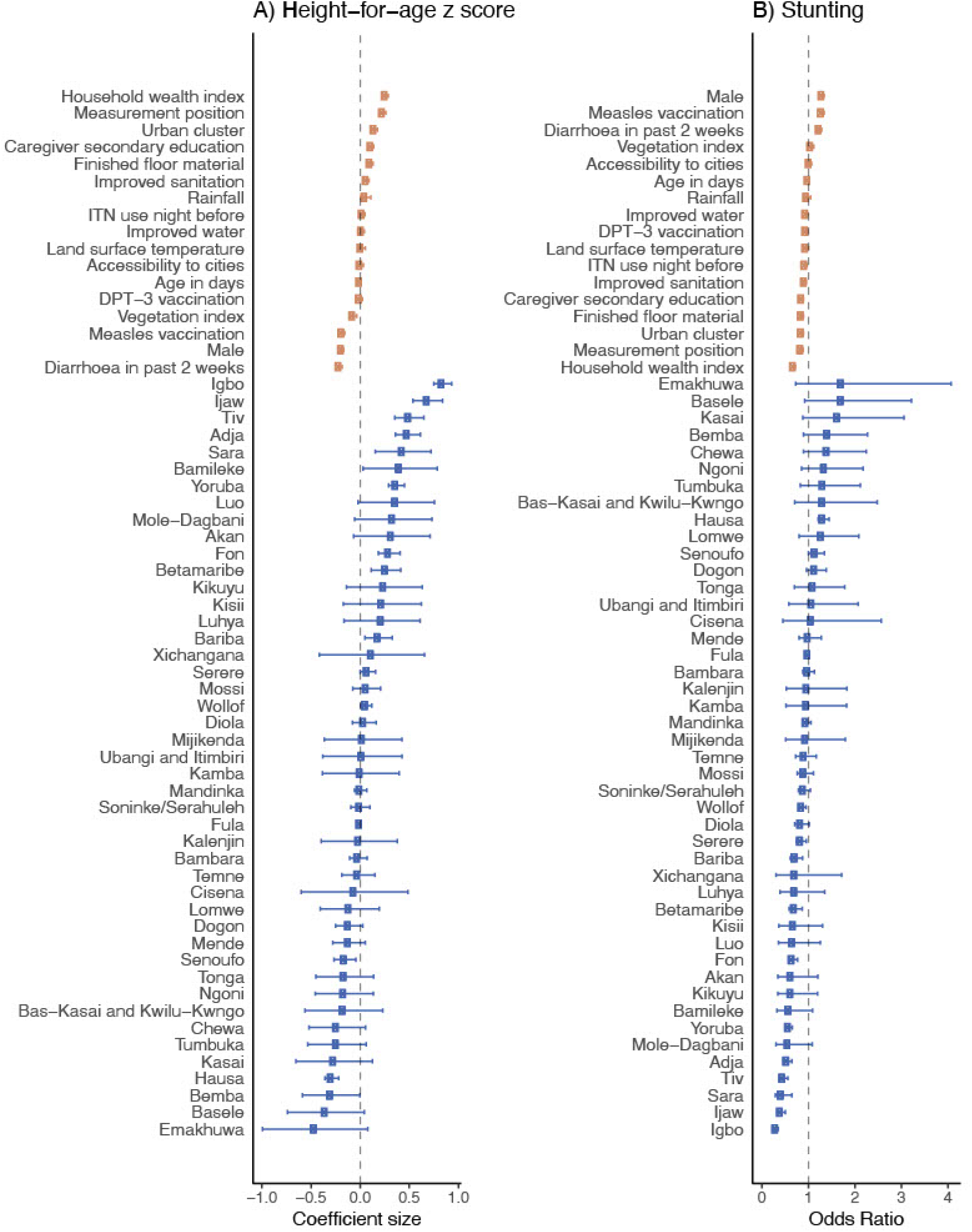
Association between ethnicity and (a) height-for-age z (HAZ) score and (b) stunting among 138,312 children aged <5 years in 18 countries in sub-Saharan Africa surveyed between 2006 to 2019. Effect estimates and 95% confidence intervals (CI) are from a generalised linear hierarchical mixed effects model controlling for all variables shown. Stunting is defined as a HAZ score more than two standard deviations lower than the World Health Organisation reference median.^36^

### Association between wasting and ethnicity

A total of 13,741 (9.9%) of 138,312 children were wasted and 58% of children had a weight-for-height lower than the WHO median. Wasting prevalence ranged from 1.1% (13 of 1,152) in Bamileke children to 21.7% (2,720 of 12,540) in Hausa children (S3 Table, S4 Table). WHZ scores differed by at least 0.5 SD in 39% of significant pairwise comparisons between ethnic groups. The largest difference between any two ethnic groups was 1.19 SD (S1 Figure). Compared to Fula children, the adjusted difference in WHZ associated with ethnicity ranged from 0.07 SD higher (95% CI 0.02-0.12, p=0.0056) in Wollof children to 1.19 SD higher (95% CI 0.97-1.42, p<0.0001) in Bamileke children. Compared to Fula children, the difference in odds of wasting associated with ethnicity ranged from 87% lower odds in Bamileke children (aOR 0.13, 95% CI 0.05-0.32, p<0.0001) to 13% lower odds in Mandinka children (aOR 0.87, 95% CI 0.76-0.99, p=0.034) (Figure 5, S5 Table). As a comparison, the association between wasting and other covariables ranged from 31% lower odds of wasting in children with height measured in a standing, compared to lying, position (aOR 0.69, 95% CI 0.65-0.73, p<0.0001) to 27% higher odds of wasting in children who had a reported episode of diarrhoea in the past two weeks compared to no episode (aOR 1.27, 95% CI 1.21-1.34, p<0.0001).

**Figure 5.**
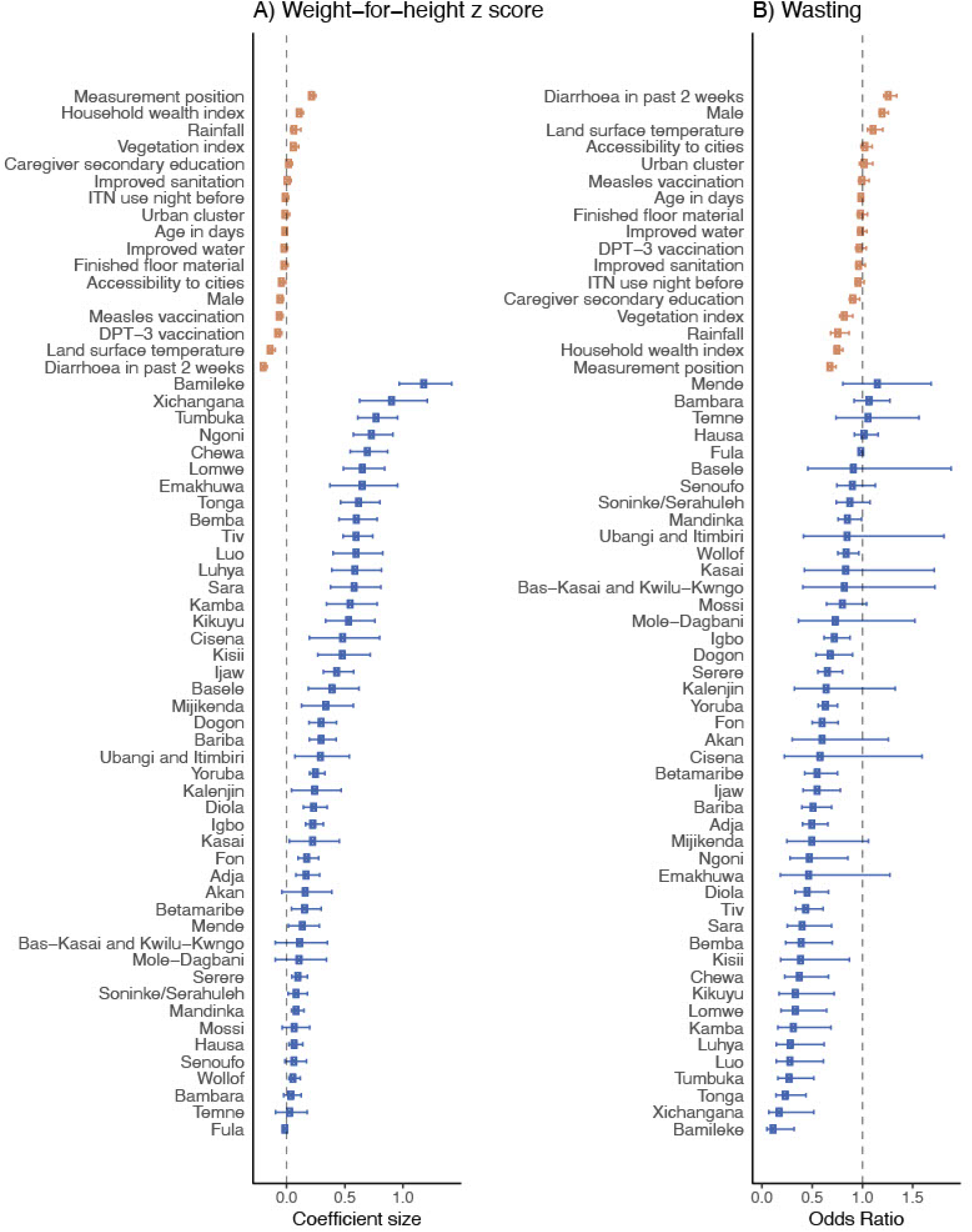
Association between ethnicity and (A) weight-for-height z (WHZ) score and (B) wasting among 138,312 children aged <5 years in 18 countries in sub-Saharan Africa surveyed between 2006 to 2019. Effect estimates and 95% confidence intervals (CI) are from a generalised linear hierarchical mixed effects model controlling for all variables shown. Wasting is defined as a WAZ score more than two standard deviations lower than the World Health Organisation reference median.^36^

### Association between underweight and ethnicity

A total of 26,297 (19.0%) of 138,312 children were underweight and 79% of children had a weight-for-height lower than the WHO median. Underweight prevalence ranged from 2.9% (33 of 1,152) in Bamileke children to 39.8% (4,994 of 12,540) in Hausa children (S3 Table, S4 Table). WAZ scores differed by at least 0.5 SD in 34% of significant pairwise comparisons between ethnic groups. The largest difference between any two ethnic groups was 1.16 SD (S1 Figure). Compared to Fula children, the adjusted difference in WAZ score associated with ethnicity ranged from 1.06 SD higher (95% CI 0.79-1.34, p<0.0001) in Bamileke children to 0.10 SD lower (95% CI -0.15, -0.04, p<0.00027) in Hausa children. Compared to Fula children, the adjusted difference in odds of underweight associated with ethnicity ranged from 85% lower odds in Bamileke children (aOR 0.15, 95% CI 0.08-0.29, p<0.0001) to 13% higher odds in Hausa children (aOR 1.13, 95% CI 1.03-1.24, p=0.010) (Figure 6, S5 Table). As a comparison, the association between underweight and other covariables ranged from 34% lower odds of underweight in wealthier households compared to less wealthy households (aOR 0.66, 95% CI 0.64-0.69, p<0.0001) to 44% higher odds of underweight in children who had a reported episode of diarrhoea in the past two weeks, compared to no episode (aOR 1.44, 95% CI 1.39-1.50, p<0.0001).

**Figure 6.**
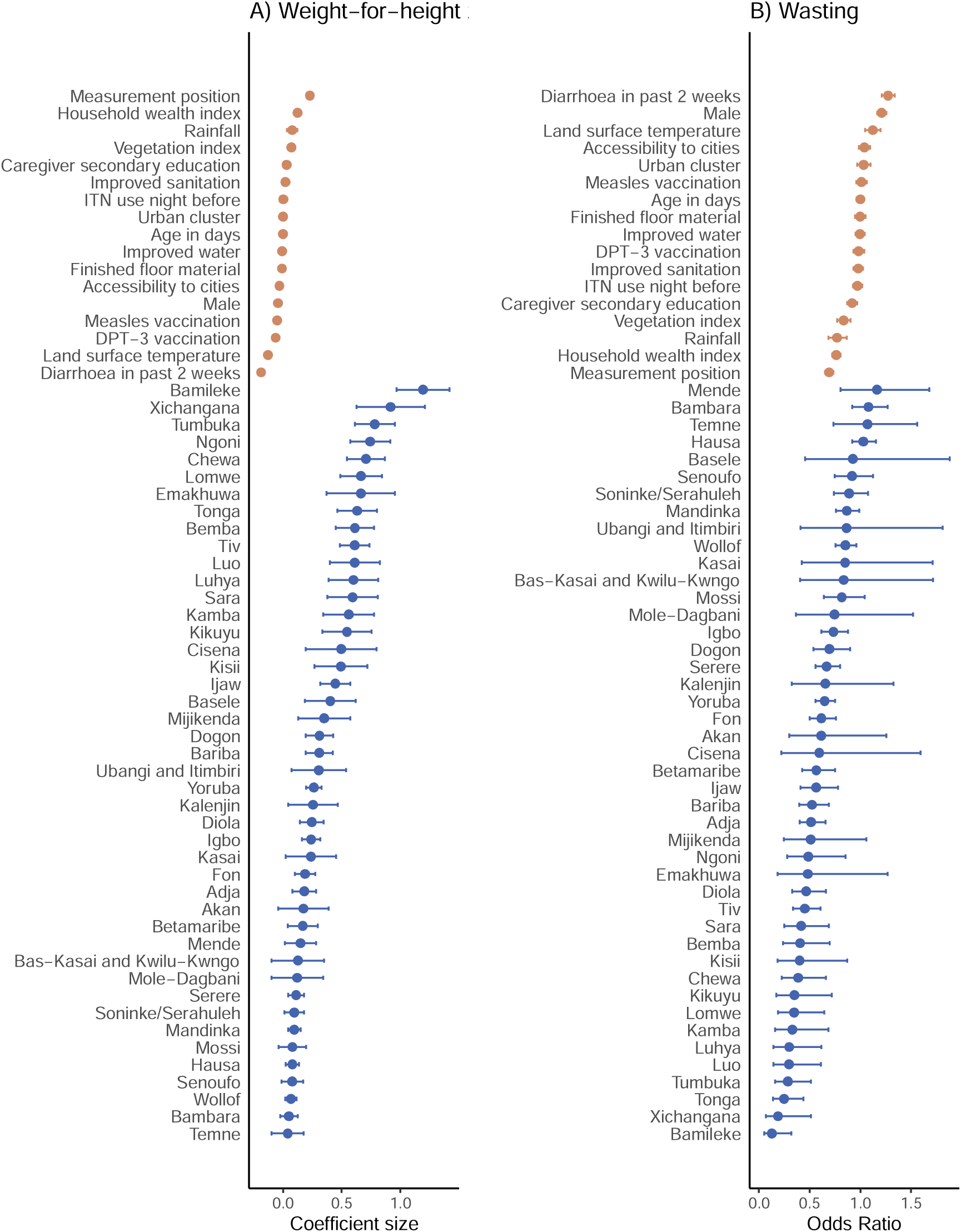
Association between ethnicity and (a) weight-for-age z (WAZ) score and (b) underweight among 138,312 children aged <5 years in 18 countries in sub-Saharan Africa surveyed between 2006 to 2019. Effect estimates and 95% confidence intervals (CI) are from a generalised linear hierarchical mixed effects model controlling for all variables shown. Underweight is defined as a WAZ score more than two standard deviations lower than the World Health Organisation reference median.^36^

### Model performance

Model performance was evaluated by calculating the area under the receiver operator characteristic curve comparing the child-level (not cluster-level) predictions for each child against the actual. For HAZ scores an AUC of 0.74 was found, for WHZ an AUC of 0.79 was found and for WHZ an AUC of 0.75 was found. These scores indicate an adequate predictive performance at the child level. Model calibration was assessed using Q-Q plots, and the plots demonstrated a close alignment with a straight line, indicating that the distribution of the model’s predictions closely matches the distribution of the observed data. This suggests that the model is well-calibrated with respect to the quantiles and distributional characteristics of the data. Predictive scores (S2-S4 Figures) indicated good performance at both child and ethnicity level. As expected, there was uncertainty in predicting growth failure in any individual child, but when examined at a population level by ethnicity this uncertainty shrunk greatly and showed a strong relationship that generalised out of sample (as measured by cross validation).

## Discussion

We investigated whether there are significant differences in anthropometric deficits in children belonging to different ethnic groups across SSA using nationally representative survey data. By analysing data for 138,312 children aged <5 years across 18 countries, we found that ethnicity is closely associated with child anthropometric deficits, accounting for differences of up to 1.30 SD in HAZ, 1.19 SD in WHZ and 1.16 SD in WAZ after adjusting for other covariables. These differences are striking, considering that an anthropometric deficit is represented by a z-score greater than 2 SD below the WHO 2006 reference median and a difference of 0.5 SD is considered clinically significant.^11^ No other covariables had associations of comparable magnitude with the malnutrition scores. Our findings suggest that ethnicity is an important factor linked to anthropometric deficits in children.

Few multi-country studies have measured differences in child growth outcomes across ethnic groups. An analysis of national survey data from Latin America found that the prevalence of stunting was on average 34% higher among indigenous children aged <5 years compared to a reference group of European or mixed ancestry children, after adjusting for wealth and place of residence (prevalence ratio 1.34, 95% CI 1.28-1.39, p=0.009).^6^ A 2014 systematic review comparing data from the WHO MGRS with data from studies performed in 55 countries or ethnic groups also found variation in height across ethnic groups.^7^ Within local studies, ethnic group differences in child growth outcomes have been found in China,^21^ Malaysia,^22^ Nepal,^23^ South Africa,^24^ the UK^25^ and the USA, but not in other studies including in Thailand^26^ and South Africa.^27^ Our study is consistent with previous studies that suggest that the burden of anthropometric deficits may be unequally distributed among some ethnic groups.

Ethnicity, a characteristic that reflects shared genetic heritage and culture, may be associated with anthropometric deficits through several environmental factors. First, ethnicity may be linked to socioeconomic risk factors for undernutrition including poor diet; inadequate water source, sanitation and hygiene; and repeat episodes of infectious disease. Overall, we found children’s height-for-age, weight-for-age and weight-for-height to be significantly lower on average among African children than the WHO median; 80% of children had HAZ scores less than zero. This finding is consistent with current evidence that body size is generally lower in LMICs than in high-income countries.^28^ Despite this, we found that ethnic group disparities persisted after adjusting for socioeconomic factors. Second, ethnicity may be linked to different cultural practices and behaviours that influence growth such as breastfeeding rates, sleep and diurnal rhythm and patterns of lifestyle and diet.^29^ For example we found growth faltering to be more common among Fula than Mandinka, Wollof and Serere children, consistent with an earlier observation that stunting is more prevalent among Fula than Mandinka children in The Gambia.^30^ The Fula have specific food cultures that may impede maternal and child nutrition; for example, pregnant Fula women do not consume important protein sources such as eggs, catfish or groundnuts.^30^ Third, ethnicity may be linked to differential access to and uptake of health care and essential services such as vaccinations. Ethnicity is a known risk factor for other child health outcomes including under five year mortality across SSA^5^ and tackling the causes of such inequalities is important to attain SDG 2.

Growth is a function of both environmental and biological potential and it is important to understand how both of these may mediate the association between ethnicity and anthropometric deficits, including the interaction between genes, hormones and the environment.^29^ For example, a recent analysis found that the odds of wasting among children were 27% higher in the hottest parts compared to the coolest parts of Africa.^16^ Population-level differences in subscapular skinfold (which affects heat loss) are also present.^31^ It is possible that growth limitation partly reflects a survival response to extreme temperatures driven by the energetic cost of thermoregulation caused by heat stress^32^ and an adaptation towards heat loss, for example through higher area-to-mass ratios.^33^ Other biological processes may be important. Sleep, for example, is not only affected by behaviour, but there is an increased occurrence of single-nucleotide polymorphisms in some African populations that reduce the duration and quality of sleep.^34^ Research is needed to understand these interconnecting vulnerabilities and their relationship with children’s growth.

The association between ethnicity and anthropometric deficits, if causative, points to the need to understand the implications for monitoring growth. Stunting and wasting, key metrics used to assess nutritional status and evaluate progress towards nutrition goals, are identified using the WHO Child Growth Standards (WHO-CGS), a universal standard based on the WHO MGRS 1997-2003 study conducted in Brazil, Ghana, India, Norway, Oman and the USA.^11^ Since identification of anthropometric deficits varies according to the growth standard used, not all countries have adopted the WHO-CGS.^35^ Such local references are more difficult to develop and implement than a universal reference, but accurate measurement of stunting and wasting is essential to ensure that anthropometric deficits are not under- or overestimated in individual children and to enable research into the putative causes and consequences of these outcomes.

Our study has several limitations. The causes of growth faltering are complicated and our analysis provides only an initial exploration of the role of ethnicity based on observational data that cannot determine causality. While we adjusted for a range of covariables, residual confounding of the relationship between ethnicity and growth outcomes by social and environmental factors is possible, and we did not adjust for nutritional intake. The measures of temperature, vegetation index and rainfall used were based on synoptic means from 2000 to 2016 which are imprecise metrics of climate conditions. Overall, we do not aim to present a comprehensive investigation, but a preliminary analysis that quantifies the relationship between ethnicity and anthropometric deficits and stimulates further work in this area.

In conclusion, significant ethnic disparities in stunting, wasting and underweight exist across SSA. Understanding and accounting for these differences is important to support effective progress monitoring and targeting of nutrition interventions.

## Contributors

LST & SB conceived of the study; LST, HSG, DJW & SB extracted and cleaned data; LST & SB verified the underlying data; SF & SWL advised on the methods; LST, SM, SF & SB conducted the analysis; LST, SF & SB wrote the first draft of the manuscript and all authors provided meaningful comments and revisions. All authors had full access to the data in the study. LST, SF & SB had final responsibility for the decision to submit for publication.

## Declaration of interests

All other authors declare no competing interests.

## Data sharing

All health data used in this analysis are available to download free of charge by registered users from the Demographic and Health Surveys Program. Registration, data, and full dataset access instructions are available online at http://dhsprogram.com. Remotely sensed data are available to download from the cited sources. Code to replicate data extraction and curation is available at: https://github.com/harry-gibson/DHS-To-Database.

## Acknowledgments

LST is a Skills Development Fellow (N011570) jointly funded by the UK Medical Research Council and the UK Department for International Development under the MRC/DFID Concordat agreement (http://www.mrc.ac.uk/). LST receives support from the Novo Nordisk Foundation (number 0069116). The authors alone are responsible for the views expressed in this Article and they do not necessarily represent the views, decisions or policies of the institutions with which they are affiliated.

## Supporting information

S1 Checklist. STROBE statement

## Competing interests

All authors declare no competing interests.

---

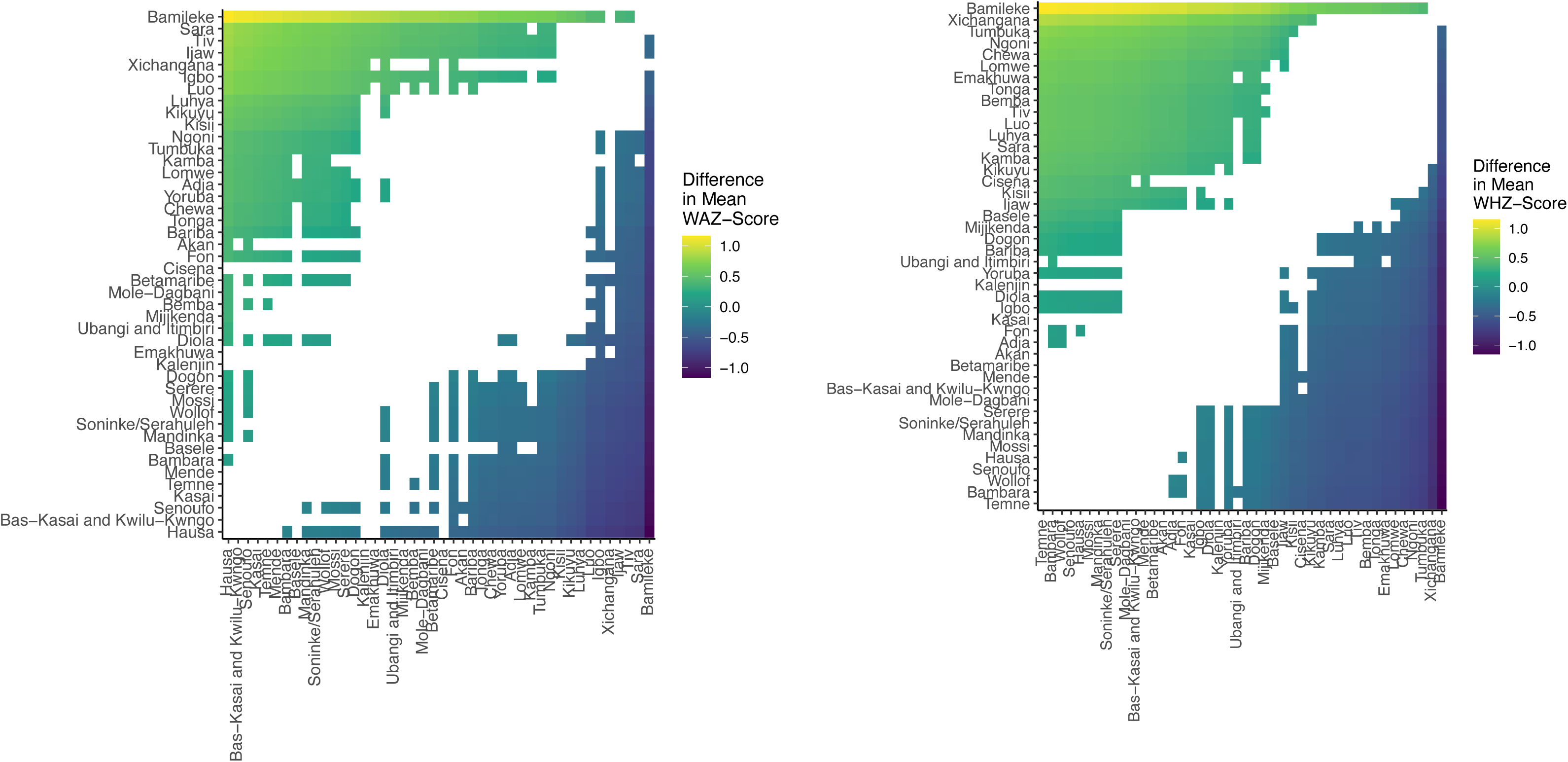

## Notes

### Competing Interest Statement

The authors have declared no competing interest.

### Author Declarations

All health data used in this analysis are available to download free of charge by registered users from the Demographic and Health Surveys Program. Registration, data, and full dataset access instructions are available online at http://dhsprogram.com. Code to replicate data extraction and curation is available at: https://github.com/harry-gibson/DHS-To-Database.

