## Supplementary material for "Ethnicity and anthropometric deficits in children: a cross-sectional analysis of national survey data from 18 countries in sub-Saharan Africa": SI

**Supplementary Information**

| **Table S1. Household- and child-level characteristics of surveys included in the analysis (n=37)** | | | | | | | | | | |  |  |  |  |
| --- | --- | --- | --- | --- | --- | --- | --- | --- | --- | --- | --- | --- | --- | --- |
| **DHS Survey** | **N** | **Household-level characteristics** | | | |  | **Child-level characteristics** | | |  |  |  |  |  |
|  |  | **Household head attended secondary education (%)** | **Urban residence (%)** | **Finished floor material (%)** | **Improved drinking water source (%)** | **Improved sanitation (%)** | **N** | **Mean age in years (%)** | **Male (%)** | **Height measured standing (not lying) (%)** | **Received DPT-3 vaccine (%)** | **Received measles-1 vaccine (%)** | **Reported ITN use previous night (%)** | **Reported diarrhoea in past two weeks (%)** |
| Benin 2012 | 4023 | 23.9 | 36.7 | 60.0 | 80.1 | 26.8 | 5589 | 2.5 | 49.8 | 72.1 | 64.4 | 66.6 | 77.2 | 7.6 |
| Benin 2017 | 4787 | 24.3 | 39.7 | 65.0 | 71.0 | 29.2 | 5975 | 1.4 | 49.4 | 30.3 | 62.5 | 47.3 | 72.2 | 14.2 |
| Burkina Faso 2010 | 2466 | 7.1 | 27.6 | 48.3 | 82.6 | 31.7 | 3873 | 2.3 | 51.6 | 61.7 | 80.9 | 72.3 | 53.4 | 14.8 |
| Cameroon 2011 | 596 | 55.0 | 73.2 | 73.2 | 82.2 | 76.7 | 926 | 2.3 | 50.1 | 49.9 | 80.2 | 75.3 | 32.9 | 13.9 |
| Cameroon 2018 | 196 | 52.6 | 62.2 | 69.9 | 78.6 | 77.6 | 245 | 1.5 | 45.7 | 35.9 | 75.9 | 63.3 | 0.0 | 12.2 |
| Chad 2014 | 1206 | 33.9 | 17.5 | 5.9 | 42.4 | 9.5 | 1886 | 2.3 | 49.2 | 52.4 | 35.3 | 56.7 | 41.3 | 26.8 |
| DRC 2007 | 1385 | 54.4 | 40.1 | 17.8 | 43.7 | 38.6 | 2099 | 2.3 | 48.3 | 53.9 | 45.1 | 56.4 | 11.8 | 16.7 |
| DRC 2013 | 3332 | 57.7 | 31.2 | 11.1 | 39.3 | 34.6 | 5479 | 2.4 | 50.0 | 55.6 | 50.9 | 59.7 | 57.9 | 17.6 |
| Cote d'Ivoire 2012 | 405 | 12.3 | 44.4 | 71.1 | 86.9 | 49.6 | 644 | 2.2 | 48.6 | 38.5 | 51.7 | 56.4 | 39.6 | 18.2 |
| Ghana 2008 | 1038 | 54.3 | 38.9 | 81.9 | 82.2 | 61.8 | 1385 | 2.4 | 50.8 | 65.1 | 80.4 | 77.8 | 44.3 | 21.6 |
| Ghana 2014 | 1255 | 57.1 | 44.7 | 92.5 | 75.1 | 66.4 | 1707 | 2.3 | 50.3 | 55.5 | 80.8 | 74.1 | 48.8 | 12.6 |
| Guinea 2012 | 1369 | 14.5 | 28.3 | 51.3 | 72.9 | 37.6 | 2150 | 2.3 | 52.5 | 58.6 | 38.4 | 51.6 | 33.2 | 18.0 |
| Guinea 2018 | 1179 | 17.8 | 27.2 | 58.7 | 74.7 | 46.7 | 1416 | 1.4 | 50.5 | 30.7 | 27.8 | 24.6 | 21.8 | 15.5 |
| Kenya 2008 | 2577 | 33.6 | 26.4 | 38.7 | 62.1 | 44.6 | 3696 | 2.4 | 50.2 | 59.6 | 76.3 | 70.8 | 55.4 | 18.1 |
| Kenya 2014 | 8923 | 36.6 | 35.7 | 40.4 | 64.5 | 46.8 | 12068 | 2.4 | 50.6 | 57.3 | 85.0 | 75.1 | 61.4 | 15.9 |
| Liberia 2013 | 76 | 28.9 | 57.9 | 40.8 | 89.5 | 55.3 | 118 | 2.4 | 50.8 | 56.8 | 75.4 | 63.6 | 50.8 | 15.3 |
| Malawi 2010 | 2267 | 23.0 | 9.9 | 18.2 | 78.1 | 14.4 | 3065 | 2.4 | 50.6 | 67.9 | 87.9 | 81.5 | 49.5 | 17.1 |
| Malawi 2015 | 2106 | 29.4 | 15.7 | 21.9 | 83.5 | 80.6 | 2267 | 1.5 | 49.9 | 35.2 | 84.1 | 70.7 | 51.3 | 28.4 |
| Mali 2006 | 3554 | 11.5 | 32.9 | 29.3 | 58.8 | 22.1 | 5339 | 2.2 | 50.8 | 60.6 | 63.9 | 60.9 | 39.2 | 13.1 |
| Mali 2012 | 2327 | 12.2 | 25.0 | 27.7 | 65.0 | 41.5 | 3612 | 2.5 | 51.0 | 61.7 | 58.5 | 63.6 | 73.8 | 8.7 |
| Mali 2018 | 500 | 19.2 | 36.4 | 52.4 | 71.6 | 57.8 | 570 | 1.4 | 46.7 | 33.0 | 59.6 | 51.2 | 0.0 | 17.0 |
| Mozambique 2011 | 2864 | 13.9 | 37.3 | 30.5 | 56.7 | 24.0 | 4219 | 2.3 | 49.6 | 57.9 | 75.6 | 74.5 | 36.4 | 10.3 |
| Nigeria 2008 | 7145 | 37.3 | 32.9 | 55.8 | 54.5 | 52.4 | 10722 | 2.3 | 49.2 | 61.2 | 33.7 | 37.8 | 7.2 | 10.0 |
| Nigeria 2013 | 9848 | 39.2 | 41.2 | 58.8 | 58.5 | 52.7 | 15458 | 2.4 | 50.1 | 53.7 | 35.7 | 36.9 | 19.8 | 10.4 |
| Nigeria 2018 | 3928 | 50.2 | 45.5 | 71.1 | 61.9 | 53.1 | 4739 | 1.5 | 51.5 | 32.7 | 47.0 | 42.8 | 0.0 | 15.2 |
| Senegal 2010 | 1314 | 8.3 | 31.8 | 54.3 | 65.9 | 43.4 | 2440 | 2.2 | 52.9 | 61.1 | 75.8 | 71.4 | 46.9 | 20.9 |
| Senegal 2012 | 2247 | 9.6 | 32.4 | 64.0 | 68.3 | 48.9 | 4682 | 2.3 | 50.7 | 60.2 | 81.0 | 69.2 | 53.1 | 15.0 |
| Senegal 2014 | 2320 | 8.6 | 32.5 | 60.1 | 70.1 | 45.9 | 4861 | 2.3 | 50.2 | 62.1 | 81.5 | 69.2 | 52.5 | 20.7 |
| Senegal 2015 | 2357 | 7.8 | 30.8 | 67.7 | 65.1 | 46.0 | 4790 | 2.3 | 50.6 | 57.8 | 80.7 | 66.6 | 58.0 | 21.7 |
| Senegal 2016 | 2355 | 9.3 | 31.8 | 69.2 | 74.6 | 50.6 | 4504 | 2.3 | 52.6 | 54.1 | 82.2 | 67.6 | 71.3 | 17.9 |
| Senegal 2019 | 2060 | 11.6 | 29.8 | 74.3 | 71.9 | 63.1 | 3196 | 1.5 | 49.8 | 33.2 | 78.6 | 62.2 | 70.2 | 16.6 |
| Sierra Leone 2008 | 886 | 23.1 | 32.4 | 32.3 | 52.0 | 40.4 | 1235 | 2.3 | 48.6 | 58.0 | 55.0 | 55.6 | 35.3 | 11.7 |
| Sierra Leone 2013 | 2052 | 22.2 | 31.6 | 37.8 | 58.2 | 49.1 | 2912 | 2.4 | 47.5 | 61.9 | 72.8 | 73.2 | 52.2 | 11.1 |
| Sierra Leone 2019 | 1573 | 30.8 | 30.7 | 47.1 | 60.5 | 48.4 | 1822 | 1.4 | 50.2 | 31.0 | 68.2 | 53.4 | 0.0 | 8.4 |
| Togo 2013 | 640 | 43.1 | 49.5 | 88.4 | 63.9 | 48.3 | 881 | 2.4 | 49.5 | 57.4 | 72.5 | 63.2 | 42.0 | 15.1 |
| Zambia 2007 | 1544 | 39.8 | 37.2 | 38.7 | 41.5 | 36.9 | 2263 | 2.4 | 49.1 | 54.3 | 34.1 | 72.8 | 34.3 | 14.8 |
| Zambia 2013 | 3888 | 47.0 | 41.6 | 41.6 | 63.1 | 44.1 | 5479 | 2.5 | 50.1 | 58.0 | 81.6 | 75.1 | 42.8 | 15.5 |
| DHS: Demographic and Health Survey; DPT: diptheria-pertussis-tetanus; DRC: Democratic Republic of the Congo; ITN: insecticide-treated net | | | | | | | | | | | | | |  |

| **Table S2. Household- and child-level characteristics of ethnic groups included in the analysis (n=45)** | | | | | | | | | |  |  |  |  |  |
| --- | --- | --- | --- | --- | --- | --- | --- | --- | --- | --- | --- | --- | --- | --- |
| **Ethnic group** | **N** | **Household-level characteristics** | | | |  | **Child-level characteristics** | | | |  |  |  |  |
|  |  | **Household head attended secondary education (%)** | **Urban residence (%)** | **Finished floor material (%)** | **Improved drinking water source (%)** | **Improved sanitation (%)** | **N** | **Mean age in years (%)** | **Male (%)** | **Height measured standing (not lying) (%)** | **Received DPT-3 vaccine (%)** | **Received measles-1 vaccine (%)** | **Reported ITN use previous night (%)** | **Reported diarrhoea in past two weeks (%)** |
| Adja | 2274 | 33.3 | 38.0 | 65.2 | 72.9 | 32.8 | 3054 | 2.2 | 50.1 | 53.2 | 64.2 | 58.9 | 67.9 | 13.4 |
| Akan | 1400 | 73.5 | 45.7 | 93.6 | 73.5 | 80.0 | 1870 | 2.4 | 49.8 | 61.9 | 81.0 | 77.2 | 47.3 | 15.0 |
| Bambara | 2389 | 12.3 | 30.8 | 29.6 | 62.9 | 31.7 | 3467 | 2.3 | 50.1 | 58.1 | 65.6 | 63.5 | 47.8 | 10.6 |
| Bamileke | 775 | 55.1 | 70.6 | 72.5 | 81.5 | 77.4 | 1152 | 2.2 | 49.1 | 47.2 | 79.9 | 73.4 | 26.5 | 13.3 |
| Bariba | 970 | 16.3 | 43.1 | 66.0 | 57.0 | 15.9 | 1307 | 1.8 | 48.8 | 43.0 | 65.7 | 56.2 | 69.0 | 14.4 |
| Bas-Kasai and Kwilu-Kwngo | 1009 | 67.0 | 37.2 | 17.3 | 33.8 | 30.6 | 1578 | 2.4 | 49.4 | 55.7 | 60.8 | 67.9 | 61.2 | 14.3 |
| Basele | 1294 | 46.0 | 30.1 | 9.6 | 56.3 | 42.5 | 2055 | 2.4 | 48.8 | 56.6 | 53.3 | 62.7 | 33.4 | 16.4 |
| Bemba | 2640 | 49.1 | 48.0 | 39.8 | 51.4 | 43.8 | 3747 | 2.4 | 49.7 | 57.4 | 69.0 | 72.8 | 42.0 | 15.2 |
| Betamaribe | 799 | 14.0 | 30.4 | 41.9 | 66.1 | 5.9 | 1117 | 1.9 | 49.4 | 52.4 | 70.9 | 59.0 | 69.8 | 16.7 |
| Chewa | 2593 | 25.6 | 16.0 | 22.1 | 74.0 | 43.1 | 3287 | 2.2 | 49.6 | 55.3 | 82.0 | 76.0 | 48.7 | 20.2 |
| Cisena | 627 | 15.8 | 28.5 | 12.3 | 49.4 | 17.4 | 1046 | 2.4 | 47.6 | 56.5 | 73.1 | 72.0 | 36.5 | 13.2 |
| Diola | 694 | 22.6 | 38.2 | 63.4 | 48.0 | 50.6 | 1015 | 2.2 | 50.4 | 56.5 | 84.8 | 74.2 | 65.3 | 19.5 |
| Dogon | 668 | 8.8 | 21.6 | 17.8 | 53.7 | 19.3 | 1009 | 2.4 | 48.8 | 61.2 | 51.4 | 57.0 | 56.2 | 9.4 |
| Emakhuwa | 1181 | 11.3 | 27.3 | 7.6 | 39.1 | 15.5 | 1711 | 2.3 | 51.2 | 60.9 | 70.0 | 72.6 | 52.3 | 8.2 |
| Fon | 3826 | 27.9 | 41.2 | 70.8 | 80.5 | 38.3 | 4841 | 2.0 | 49.5 | 55.2 | 65.6 | 60.5 | 78.6 | 7.9 |
| Fula | 11172 | 10.0 | 25.3 | 45.9 | 58.0 | 36.1 | 17558 | 2.1 | 50.5 | 53.2 | 52.7 | 48.4 | 42.4 | 17.8 |
| Hausa | 8012 | 24.3 | 25.6 | 42.9 | 54.9 | 56.7 | 12540 | 2.2 | 49.2 | 53.3 | 14.3 | 21.6 | 11.5 | 13.5 |
| Igbo | 3951 | 51.5 | 59.8 | 81.0 | 68.2 | 61.4 | 5752 | 2.2 | 51.0 | 54.5 | 71.6 | 61.9 | 16.5 | 7.4 |
| Ijaw | 1066 | 65.8 | 22.9 | 72.7 | 32.0 | 14.8 | 1554 | 2.2 | 50.8 | 48.5 | 40.9 | 45.9 | 17.0 | 2.8 |
| Kalenjin | 2254 | 32.3 | 19.0 | 25.1 | 46.8 | 40.2 | 3251 | 2.4 | 51.1 | 55.5 | 84.7 | 72.6 | 50.8 | 12.5 |
| Kamba | 1336 | 31.4 | 37.4 | 49.3 | 53.0 | 50.1 | 1765 | 2.4 | 50.7 | 56.5 | 86.7 | 77.0 | 58.9 | 15.1 |
| Kasai | 1763 | 59.6 | 38.2 | 15.4 | 40.0 | 31.5 | 2881 | 2.3 | 49.3 | 53.5 | 45.8 | 52.1 | 39.7 | 20.6 |
| Kikuyu | 2232 | 47.0 | 44.2 | 57.0 | 76.5 | 58.6 | 2664 | 2.5 | 51.5 | 58.4 | 88.2 | 80.8 | 48.5 | 10.4 |
| Kisii | 870 | 49.1 | 31.8 | 36.1 | 74.6 | 41.7 | 1126 | 2.5 | 51.8 | 62.2 | 86.4 | 77.2 | 69.4 | 11.7 |
| Lomwe | 972 | 24.1 | 14.7 | 19.7 | 86.9 | 47.5 | 1151 | 2.0 | 50.5 | 52.2 | 87.6 | 77.9 | 48.7 | 23.6 |
| Luhya | 2039 | 35.8 | 33.1 | 33.3 | 76.3 | 38.9 | 2862 | 2.4 | 50.5 | 59.1 | 81.6 | 72.7 | 68.0 | 20.5 |
| Luo | 1782 | 34.1 | 39.8 | 45.7 | 59.1 | 49.2 | 2556 | 2.4 | 49.8 | 58.1 | 74.7 | 69.0 | 67.9 | 22.5 |
| Mandinka | 3108 | 17.5 | 36.0 | 54.3 | 68.5 | 42.7 | 5238 | 2.1 | 51.4 | 53.5 | 61.7 | 57.5 | 49.1 | 18.2 |
| Mende | 2315 | 25.4 | 25.8 | 33.8 | 59.9 | 41.1 | 3025 | 2.1 | 48.9 | 52.6 | 72.4 | 67.3 | 37.3 | 8.2 |
| Mijikenda | 987 | 17.2 | 29.8 | 30.1 | 63.3 | 41.4 | 1540 | 2.4 | 48.1 | 57.3 | 79.8 | 71.2 | 66.0 | 22.4 |
| Mole-Dagbani | 893 | 28.1 | 36.4 | 78.4 | 85.8 | 39.6 | 1222 | 2.3 | 51.6 | 56.5 | 80.0 | 73.6 | 46.0 | 19.1 |
| Mossi | 1889 | 7.5 | 28.8 | 53.1 | 83.2 | 34.3 | 2981 | 2.3 | 52.1 | 62.8 | 83.5 | 73.9 | 54.8 | 14.3 |
| Ngoni | 1087 | 31.5 | 27.5 | 32.8 | 79.2 | 49.9 | 1359 | 2.2 | 51.4 | 55.2 | 81.3 | 79.4 | 48.1 | 17.2 |
| Sara | 1156 | 35.2 | 17.6 | 6.5 | 41.9 | 9.5 | 1803 | 2.3 | 49.3 | 52.4 | 36.4 | 57.7 | 40.8 | 27.0 |
| Senoufo | 1144 | 11.6 | 29.2 | 41.3 | 70.0 | 28.8 | 1846 | 2.2 | 52.2 | 52.7 | 64.5 | 63.9 | 49.1 | 16.2 |
| Serere | 1674 | 10.0 | 30.2 | 66.1 | 69.9 | 53.0 | 3228 | 2.3 | 51.5 | 59.0 | 83.4 | 70.6 | 59.4 | 17.1 |
| Soninke/Serahuleh | 1148 | 10.6 | 37.2 | 44.9 | 66.5 | 42.3 | 1959 | 2.3 | 49.1 | 61.7 | 65.8 | 64.3 | 52.2 | 13.7 |
| Temne | 1947 | 26.0 | 34.6 | 43.0 | 54.7 | 51.5 | 2655 | 2.0 | 48.0 | 49.8 | 62.8 | 58.7 | 28.5 | 12.7 |
| Tiv | 768 | 55.1 | 11.5 | 43.5 | 49.3 | 22.7 | 1176 | 2.2 | 50.3 | 52.7 | 26.7 | 32.7 | 9.3 | 8.8 |
| Tonga | 1468 | 42.0 | 26.0 | 39.8 | 62.7 | 39.2 | 2134 | 2.4 | 49.1 | 55.1 | 65.7 | 75.1 | 38.6 | 16.9 |
| Tumbuka | 1045 | 40.9 | 23.3 | 32.1 | 69.0 | 43.7 | 1396 | 2.2 | 51.6 | 56.7 | 75.1 | 75.7 | 42.3 | 17.8 |
| Ubangi and Itimbiri | 651 | 54.2 | 23.7 | 7.1 | 21.5 | 41.8 | 1064 | 2.4 | 51.8 | 56.1 | 34.0 | 55.8 | 58.6 | 14.7 |
| Wollof | 4161 | 7.7 | 33.0 | 75.8 | 83.4 | 65.0 | 9089 | 2.2 | 51.2 | 54.6 | 83.0 | 70.0 | 56.7 | 17.8 |
| Xichangana | 1056 | 15.6 | 53.7 | 66.9 | 80.7 | 37.5 | 1462 | 2.3 | 49.2 | 55.4 | 84.0 | 78.5 | 17.9 | 10.7 |
| Yoruba | 5503 | 60.2 | 63.2 | 86.4 | 76.3 | 55.8 | 7219 | 2.2 | 50.7 | 51.8 | 67.2 | 61.1 | 22.8 | 7.7 |
| DPT: diptheria-pertussis-tetanus; DRC: Democratic Republic of the Congo; ITN: insecticide-treated net | | | | | | | | | |  |  |  |  |  |

| **Table S3. Anthropometric characteristics for surveys included in the analysis (n=37)** | | | | | | | |  | |
| --- | --- | --- | --- | --- | --- | --- | --- | --- | --- |
| **DHS Survey** | **N** | **Height in cm (median, N)** | **HAZ score (median, N)** | **Stunted (%, N)** | **WHZ score (median, N)** | **Wasted (%, N)** | **WAZ score (median, N)** | | **Underweight (%, N)** |
| Benin 2012 | 5589 | 83.0 (5589) | -1.62 (5589) | 42.5 (5589) | -0.02 (5589) | 15.8 (5589) | -0.93 (5589) | | 20.2 (5589) |
| Benin 2017 | 5975 | 75.4 (5975) | -1.37 (5975) | 30.8 (5975) | -0.30 (5975) | 6.3 (5975) | -0.98 (5975) | | 17.3 (5975) |
| Burkina Faso 2010 | 3873 | 83.0 (3873) | -1.41 (3873) | 33.7 (3873) | -0.63 (3873) | 17.0 (3873) | -1.21 (3873) | | 25.4 (3873) |
| Cameroon 2011 | 926 | 84.2 (926) | -0.99 (926) | 22.5 (926) | 0.78 (926) | 1.2 (926) | 0.04 (926) | | 2.9 (926) |
| Cameroon 2018 | 245 | 78.2 (245) | -0.70 (245) | 18.0 (245) | 0.99 (245) | 1.2 (245) | 0.38 (245) | | 3.7 (245) |
| Chad 2014 | 1886 | 83.0 (1886) | -1.13 (1886) | 29.7 (1886) | -0.17 (1886) | 9.6 (1886) | -0.78 (1886) | | 17.7 (1886) |
| DRC 2007 | 2099 | 82.0 (2099) | -1.78 (2099) | 44.9 (2099) | -0.13 (2099) | 9.5 (2099) | -1.00 (2099) | | 22.0 (2099) |
| DRC 2013 | 5479 | 81.9 (5479) | -1.79 (5479) | 45.6 (5479) | -0.14 (5479) | 7.6 (5479) | -1.10 (5479) | | 23.7 (5479) |
| Cote d'Ivoire 2012 | 644 | 81.6 (644) | -1.22 (644) | 29.5 (644) | -0.23 (644) | 5.6 (644) | -0.88 (644) | | 13.0 (644) |
| Ghana 2008 | 1385 | 84.2 (1385) | -1.21 (1385) | 28.7 (1385) | -0.37 (1385) | 9.1 (1385) | -0.89 (1385) | | 14.8 (1385) |
| Ghana 2014 | 1707 | 84.5 (1707) | -0.96 (1707) | 18.6 (1707) | -0.30 (1707) | 5.5 (1707) | -0.75 (1707) | | 11.4 (1707) |
| Guinea 2012 | 2150 | 83.0 (2150) | -1.22 (2150) | 31.4 (2150) | -0.41 (2150) | 11.5 (2150) | -0.91 (2150) | | 19.3 (2150) |
| Guinea 2018 | 1416 | 75.3 (1416) | -1.03 (1416) | 27.6 (1416) | -0.34 (1416) | 10.1 (1416) | -0.81 (1416) | | 17.1 (1416) |
| Kenya 2008 | 3696 | 83.5 (3696) | -1.46 (3696) | 34.7 (3696) | 0.01 (3696) | 5.8 (3696) | -0.78 (3696) | | 14.8 (3696) |
| Kenya 2014 | 12068 | 85.0 (12068) | -1.25 (12068) | 27.5 (12068) | 0.06 (12068) | 3.3 (12068) | -0.66 (12068) | | 10.6 (12068) |
| Liberia 2013 | 118 | 85.2 (118) | -1.32 (118) | 26.3 (118) | -0.29 (118) | 4.2 (118) | -0.92 (118) | | 11.0 (118) |
| Malawi 2010 | 3065 | 83.0 (3065) | -1.88 (3065) | 46.6 (3065) | 0.34 (3065) | 3.8 (3065) | -0.75 (3065) | | 12.3 (3065) |
| Malawi 2015 | 2267 | 76.5 (2267) | -1.52 (2267) | 34.7 (2267) | 0.15 (2267) | 3.5 (2267) | -0.70 (2267) | | 10.8 (2267) |
| Mali 2006 | 5339 | 80.0 (5339) | -1.40 (5339) | 35.7 (5339) | -0.56 (5339) | 16.3 (5339) | -1.16 (5339) | | 25.6 (5339) |
| Mali 2012 | 3612 | 84.3 (3612) | -1.47 (3612) | 38.3 (3612) | -0.47 (3612) | 12.8 (3612) | -1.16 (3612) | | 25.9 (3612) |
| Mali 2018 | 570 | 76.8 (570) | -1.04 (570) | 24.9 (570) | -0.55 (570) | 12.3 (570) | -0.96 (570) | | 20.2 (570) |
| Mozambique 2011 | 4219 | 82.3 (4219) | -1.67 (4219) | 39.6 (4219) | 0.27 (4219) | 5.0 (4219) | -0.76 (4219) | | 14.3 (4219) |
| Nigeria 2008 | 10722 | 82.2 (10722) | -1.54 (10722) | 40.4 (10722) | -0.18 (10722) | 15.9 (10722) | -1.02 (10722) | | 24.9 (10722) |
| Nigeria 2013 | 15458 | 82.4 (15458) | -1.43 (15458) | 38.1 (15458) | -0.56 (15458) | 17.9 (15458) | -1.24 (15458) | | 29.4 (15458) |
| Nigeria 2018 | 4739 | 75.5 (4739) | -1.42 (4739) | 35.3 (4739) | -0.36 (4739) | 9.1 (4739) | -1.07 (4739) | | 23.5 (4739) |
| Senegal 2010 | 2440 | 82.6 (2440) | -1.21 (2440) | 28.2 (2440) | -0.53 (2440) | 10.3 (2440) | -1.04 (2440) | | 19.3 (2440) |
| Senegal 2012 | 4682 | 84.1 (4682) | -0.95 (4682) | 18.7 (4682) | -0.65 (4682) | 10.2 (4682) | -0.96 (4682) | | 16.8 (4682) |
| Senegal 2014 | 4861 | 84.2 (4861) | -1.01 (4861) | 20.1 (4861) | -0.47 (4861) | 6.7 (4861) | -0.88 (4861) | | 13.9 (4861) |
| Senegal 2015 | 4790 | 84.0 (4790) | -1.07 (4790) | 20.8 (4790) | -0.56 (4790) | 7.8 (4790) | -0.99 (4790) | | 16.4 (4790) |
| Senegal 2016 | 4504 | 83.4 (4504) | -0.95 (4504) | 17.1 (4504) | -0.54 (4504) | 7.1 (4504) | -0.91 (4504) | | 14.3 (4504) |
| Senegal 2019 | 3196 | 76.8 (3196) | -1.03 (3196) | 20.9 (3196) | -0.47 (3196) | 8.4 (3196) | -0.88 (3196) | | 15.5 (3196) |
| Sierra Leone 2008 | 1235 | 81.4 (1235) | -1.39 (1235) | 35.9 (1235) | -0.22 (1235) | 11.4 (1235) | -0.85 (1235) | | 21.0 (1235) |
| Sierra Leone 2013 | 2912 | 83.0 (2912) | -1.51 (2912) | 37.6 (2912) | 0.10 (2912) | 8.8 (2912) | -0.79 (2912) | | 15.1 (2912) |
| Sierra Leone 2019 | 1822 | 75.3 (1822) | -1.27 (1822) | 30.1 (1822) | -0.06 (1822) | 6.8 (1822) | -0.76 (1822) | | 15.4 (1822) |
| Togo 2013 | 881 | 84.6 (881) | -1.14 (881) | 25.3 (881) | -0.22 (881) | 5.9 (881) | -0.79 (881) | | 13.4 (881) |
| Zambia 2007 | 2263 | 82.0 (2263) | -1.82 (2263) | 44.9 (2263) | 0.33 (2263) | 4.6 (2263) | -0.76 (2263) | | 13.1 (2263) |
| Zambia 2013 | 5479 | 83.5 (5479) | -1.70 (5479) | 41.0 (5479) | 0.03 (5479) | 6.1 (5479) | -0.93 (5479) | | 15.0 (5479) |
| DHS: Demographic and Health Survey; DRC: Democratic Republic of the Congo; HAZ: height-for-age z score; WAZ: weight-for-age z score; WHZ: weight-for-height z score | | | | | | | | | |

| **Table S4. Anthropometric characteristics for ethnic groups included in the analysis (n=45)** | | | | | | | | |
| --- | --- | --- | --- | --- | --- | --- | --- | --- |
| **Ethnic group** | **N** | **Height in cm (median, N)** | **HAZ score (median, N)** | **Stunted (%, N)** | **WHZ score (median, N)** | **Wasted (%, N)** | **WAZ score (median, N)** | **Underweight (%, N)** |
| Adja | 3054 | 81.4 (3054) | -1.27 (3054) | 31.5 (3054) | -0.21 (3054) | 8.9 (3054) | -0.87 (3054) | 16.1 (3054) |
| Akan | 1870 | 85.3 (1870) | -0.99 (1870) | 22.6 (1870) | -0.25 (1870) | 5.6 (1870) | -0.73 (1870) | 11.1 (1870) |
| Bambara | 3467 | 81.2 (3467) | -1.41 (3467) | 36.3 (3467) | -0.51 (3467) | 16.0 (3467) | -1.16 (3467) | 25.1 (3467) |
| Bamileke | 1152 | 82.5 (1152) | -0.90 (1152) | 21.5 (1152) | 0.80 (1152) | 1.1 (1152) | 0.12 (1152) | 2.9 (1152) |
| Bariba | 1307 | 77.2 (1307) | -1.57 (1307) | 36.8 (1307) | -0.17 (1307) | 9.1 (1307) | -0.91 (1307) | 18.2 (1307) |
| Bas-Kasai and Kwilu-Kwngo | 1578 | 82.7 (1578) | -1.70 (1578) | 41.6 (1578) | -0.39 (1578) | 7.8 (1578) | -1.20 (1578) | 24.8 (1578) |
| Basele | 2055 | 81.5 (2055) | -1.98 (2055) | 49.7 (2055) | 0.05 (2055) | 8.3 (2055) | -1.05 (2055) | 23.1 (2055) |
| Bemba | 3747 | 82.4 (3747) | -1.79 (3747) | 43.7 (3747) | 0.07 (3747) | 6.3 (3747) | -0.95 (3747) | 16.7 (3747) |
| Betamaribe | 1117 | 76.8 (1117) | -1.68 (1117) | 42.4 (1117) | -0.37 (1117) | 12.5 (1117) | -1.15 (1117) | 23.2 (1117) |
| Chewa | 3287 | 79.9 (3287) | -1.78 (3287) | 43.7 (3287) | 0.23 (3287) | 4.6 (3287) | -0.78 (3287) | 12.0 (3287) |
| Cisena | 1046 | 83.6 (1046) | -1.73 (1046) | 40.0 (1046) | -0.05 (1046) | 7.4 (1046) | -0.93 (1046) | 16.7 (1046) |
| Diola | 1015 | 82.8 (1015) | -1.00 (1015) | 20.2 (1015) | -0.22 (1015) | 4.0 (1015) | -0.72 (1015) | 9.5 (1015) |
| Dogon | 1009 | 81.6 (1009) | -1.58 (1009) | 41.0 (1009) | -0.42 (1009) | 12.4 (1009) | -1.10 (1009) | 26.1 (1009) |
| Emakhuwa | 1711 | 81.0 (1711) | -2.07 (1711) | 51.7 (1711) | 0.21 (1711) | 6.0 (1711) | -1.05 (1711) | 18.8 (1711) |
| Fon | 4841 | 79.0 (4841) | -1.43 (4841) | 35.6 (4841) | -0.16 (4841) | 11.0 (4841) | -0.91 (4841) | 17.6 (4841) |
| Fula | 17558 | 80.4 (17558) | -1.28 (17558) | 32.1 (17558) | -0.62 (17558) | 13.9 (17558) | -1.17 (17558) | 25.2 (17558) |
| Hausa | 12540 | 78.5 (12540) | -2.12 (12540) | 52.3 (12540) | -0.60 (12540) | 21.7 (12540) | -1.65 (12540) | 39.8 (12540) |
| Igbo | 5752 | 83.3 (5752) | -0.64 (5752) | 17.8 (5752) | -0.26 (5752) | 9.4 (5752) | -0.54 (5752) | 10.6 (5752) |
| Ijaw | 1554 | 83.2 (1554) | -1.06 (1554) | 26.4 (1554) | 0.00 (1554) | 6.8 (1554) | -0.55 (1554) | 12.1 (1554) |
| Kalenjin | 3251 | 84.1 (3251) | -1.53 (3251) | 35.2 (3251) | -0.28 (3251) | 5.9 (3251) | -1.01 (3251) | 18.4 (3251) |
| Kamba | 1765 | 83.8 (1765) | -1.46 (1765) | 32.9 (1765) | 0.03 (1765) | 3.6 (1765) | -0.79 (1765) | 12.9 (1765) |
| Kasai | 2881 | 81.9 (2881) | -1.81 (2881) | 46.3 (2881) | -0.18 (2881) | 8.4 (2881) | -1.09 (2881) | 24.2 (2881) |
| Kikuyu | 2664 | 86.7 (2664) | -1.12 (2664) | 22.3 (2664) | 0.14 (2664) | 3.1 (2664) | -0.50 (2664) | 7.2 (2664) |
| Kisii | 1126 | 85.0 (1126) | -1.25 (1126) | 26.1 (1126) | 0.16 (1126) | 3.1 (1126) | -0.57 (1126) | 10.1 (1126) |
| Lomwe | 1151 | 79.6 (1151) | -1.68 (1151) | 40.0 (1151) | 0.21 (1151) | 3.4 (1151) | -0.72 (1151) | 11.1 (1151) |
| Luhya | 2862 | 84.6 (2862) | -1.18 (2862) | 27.0 (2862) | 0.22 (2862) | 2.5 (2862) | -0.54 (2862) | 8.3 (2862) |
| Luo | 2556 | 84.4 (2556) | -1.07 (2556) | 25.3 (2556) | 0.19 (2556) | 2.9 (2556) | -0.43 (2556) | 8.0 (2556) |
| Mandinka | 5238 | 81.3 (5238) | -1.15 (5238) | 26.9 (5238) | -0.39 (5238) | 9.1 (5238) | -0.89 (5238) | 17.2 (5238) |
| Mende | 3025 | 79.7 (3025) | -1.51 (3025) | 37.3 (3025) | 0.03 (3025) | 8.9 (3025) | -0.83 (3025) | 16.8 (3025) |
| Mijikenda | 1540 | 82.9 (1540) | -1.57 (1540) | 36.4 (1540) | -0.17 (1540) | 6.0 (1540) | -0.95 (1540) | 16.7 (1540) |
| Mole-Dagbani | 1222 | 83.1 (1222) | -1.13 (1222) | 24.0 (1222) | -0.46 (1222) | 9.4 (1222) | -0.96 (1222) | 15.7 (1222) |
| Mossi | 2981 | 83.1 (2981) | -1.37 (2981) | 32.5 (2981) | -0.63 (2981) | 16.7 (2981) | -1.19 (2981) | 24.2 (2981) |
| Ngoni | 1359 | 81.9 (1359) | -1.72 (1359) | 41.6 (1359) | 0.28 (1359) | 5.7 (1359) | -0.72 (1359) | 12.4 (1359) |
| Sara | 1803 | 83.0 (1803) | -1.12 (1803) | 28.6 (1803) | -0.12 (1803) | 8.8 (1803) | -0.74 (1803) | 16.5 (1803) |
| Senoufo | 1846 | 81.0 (1846) | -1.55 (1846) | 38.9 (1846) | -0.45 (1846) | 12.7 (1846) | -1.22 (1846) | 25.6 (1846) |
| Serere | 3228 | 83.8 (3228) | -0.98 (3228) | 18.8 (3228) | -0.49 (3228) | 6.4 (3228) | -0.88 (3228) | 13.0 (3228) |
| Soninke/Serahuleh | 1959 | 82.1 (1959) | -1.16 (1959) | 28.8 (1959) | -0.58 (1959) | 13.4 (1959) | -1.07 (1959) | 21.6 (1959) |
| Temne | 2655 | 79.3 (2655) | -1.29 (2655) | 32.5 (2655) | -0.05 (2655) | 8.6 (2655) | -0.78 (2655) | 16.1 (2655) |
| Tiv | 1176 | 81.5 (1176) | -1.31 (1176) | 31.7 (1176) | 0.09 (1176) | 7.8 (1176) | -0.63 (1176) | 12.8 (1176) |
| Tonga | 2134 | 82.5 (2134) | -1.60 (2134) | 37.8 (2134) | 0.06 (2134) | 3.9 (2134) | -0.85 (2134) | 12.4 (2134) |
| Tumbuka | 1396 | 81.2 (1396) | -1.69 (1396) | 41.3 (1396) | 0.32 (1396) | 3.6 (1396) | -0.69 (1396) | 11.7 (1396) |
| Ubangi and Itimbiri | 1064 | 81.6 (1064) | -1.60 (1064) | 40.4 (1064) | -0.06 (1064) | 7.4 (1064) | -0.93 (1064) | 18.1 (1064) |
| Wollof | 9089 | 82.7 (9089) | -0.94 (9089) | 18.4 (9089) | -0.53 (9089) | 8.0 (9089) | -0.89 (9089) | 14.0 (9089) |
| Xichangana | 1462 | 83.2 (1462) | -1.22 (1462) | 25.3 (1462) | 0.47 (1462) | 2.3 (1462) | -0.34 (1462) | 7.3 (1462) |
| Yoruba | 7219 | 81.9 (7219) | -1.14 (7219) | 27.9 (7219) | -0.24 (7219) | 9.6 (7219) | -0.80 (7219) | 14.6 (7219) |
| DRC: Democratic Republic of the Congo; HAZ: height-for-age z score; WAZ: weight-for-age z score; WHZ: weight-for-height z score | | | | | | | | |

| **Table S5. Association between ethnicity and growth outcomes in children (n=138,312) aged <5 years in sub-Saharan Africa** | | | | | | | | | | | | |
| --- | --- | --- | --- | --- | --- | --- | --- | --- | --- | --- | --- | --- |
|  | **Stunting** | | **Wasting** | | **Underweight** | | **HAZ score** | | **WHZ score** | | **WAZ score** | |
|  | OR (95% CI) | p | OR (95% CI) | p | OR (95% CI) | p | Coefficient (95% CI) | p | Coefficient (95% CI) | p | Coefficient (95% CI) | p |
| ***Ethnic group*** |  |  |  |  |  |  |  |  |  |  |  |  |
| Adja | 0.55 (0.46, 0.64) | <0.0001 | 0.51 (0.40, 0.66) | <0.0001 | 0.53 (0.43, 0.64) | <0.0001 | 0.49 (0.36, 0.61) | <0.0001 | 0.18 (0.08, 0.28) | 0.00044 | 0.37 (0.28, 0.47) | <0.0001 |
| Akan | 0.64 (0.34, 1.20) | 0.16 | 0.62 (0.30, 1.26) | 0.18 | 0.55 (0.33, 0.91) | 0.020 | 0.32 (-0.07, 0.71) | 0.10 | 0.17 (-0.04, 0.39) | 0.11 | 0.29 (0.01, 0.57) | 0.041 |
| Bambara | 1.00 (0.88, 1.13) | 0.95 | 1.08 (0.92, 1.27) | 0.36 | 0.87 (0.76, 1.00) | 0.048 | -0.02 (-0.11, 0.07) | 0.68 | 0.05 (-0.02, 0.12) | 0.19 | 0.03 (-0.04, 0.10) | 0.38 |
| Bamileke | 0.59 (0.32, 1.09) | 0.092 | 0.13 (0.05, 0.32) | <0.0001 | 0.15 (0.08, 0.29) | <0.0001 | 0.41 (0.03, 0.78) | 0.036 | 1.19 (0.97, 1.42) | <0.0001 | 1.06 (0.79, 1.34) | <0.0001 |
| Bariba | 0.73 (0.61, 0.87) | 0.00053 | 0.52 (0.40, 0.69) | <0.0001 | 0.59 (0.48, 0.73) | <0.0001 | 0.19 (0.05, 0.33) | 0.0085 | 0.31 (0.19, 0.42) | <0.0001 | 0.30 (0.19, 0.40) | <0.0001 |
| Bas-Kasai and Kwilu-Kwngo | 1.32 (0.70, 2.48) | 0.39 | 0.84 (0.41, 1.72) | 0.63 | 1.14 (0.69, 1.91) | 0.61 | -0.17 (-0.56, 0.23) | 0.41 | 0.13 (-0.10, 0.35) | 0.27 | -0.05 (-0.33, 0.24) | 0.73 |
| Basele | 1.72 (0.92, 3.21) | 0.092 | 0.93 (0.46, 1.88) | 0.83 | 1.07 (0.64, 1.77) | 0.81 | -0.35 (-0.74, 0.04) | 0.08 | 0.40 (0.19, 0.62) | 0.00028 | 0.04 (-0.24, 0.32) | 0.78 |
| Bemba | 1.42 (0.89, 2.27) | 0.14 | 0.41 (0.24, 0.70) | 0.0011 | 0.63 (0.43, 0.93) | 0.019 | -0.30 (-0.59, -0.00) | 0.047 | 0.61 (0.45, 0.78) | <0.0001 | 0.25 (0.04, 0.45) | 0.022 |
| Betamaribe | 0.71 (0.59, 0.86) | 0.00055 | 0.57 (0.43, 0.75) | <0.0001 | 0.60 (0.48, 0.75) | <0.0001 | 0.26 (0.11, 0.41) | 0.00069 | 0.17 (0.04, 0.30) | 0.0094 | 0.25 (0.14, 0.37) | <0.0001 |
| Chewa | 1.41 (0.89, 2.24) | 0.14 | 0.39 (0.23, 0.66) | 0.00052 | 0.44 (0.30, 0.64) | <0.0001 | -0.23 (-0.52, 0.05) | 0.11 | 0.71 (0.55, 0.87) | <0.0001 | 0.35 (0.14, 0.56) | 0.00091 |
| Cisena | 1.07 (0.45, 2.56) | 0.87 | 0.59 (0.22, 1.59) | 0.30 | 0.59 (0.29, 1.18) | 0.14 | -0.06 (-0.60, 0.49) | 0.84 | 0.50 (0.19, 0.80) | 0.0013 | 0.28 (-0.11, 0.67) | 0.17 |
| Diola | 0.84 (0.70, 1.01) | 0.071 | 0.47 (0.33, 0.66) | <0.0001 | 0.55 (0.43, 0.69) | <0.0001 | 0.04 (-0.08, 0.16) | 0.50 | 0.25 (0.14, 0.35) | <0.0001 | 0.19 (0.10, 0.28) | <0.0001 |
| Dogon | 1.15 (0.96, 1.38) | 0.13 | 0.70 (0.54, 0.90) | 0.0059 | 0.76 (0.62, 0.93) | 0.0080 | -0.11 (-0.25, 0.03) | 0.11 | 0.31 (0.19, 0.43) | <0.0001 | 0.15 (0.04, 0.25) | 0.0058 |
| Emakhuwa | 1.72 (0.73, 4.07) | 0.22 | 0.48 (0.18, 1.27) | 0.14 | 0.66 (0.33, 1.31) | 0.23 | -0.46 (-0.99, 0.07) | 0.092 | 0.66 (0.37, 0.95) | <0.0001 | 0.17 (-0.22, 0.55) | 0.39 |
| Fon | 0.66 (0.57, 0.76) | <0.0001 | 0.62 (0.50, 0.76) | <0.0001 | 0.61 (0.52, 0.72) | <0.0001 | 0.30 (0.19, 0.41) | <0.0001 | 0.19 (0.10, 0.28) | <0.0001 | 0.28 (0.20, 0.37) | <0.0001 |
| Fula | Reference | - | Reference | - | Reference | - | Reference | - | Reference | - | Reference | - |
| Hausa | 1.32 (1.21, 1.44) | <0.0001 | 1.03 (0.92, 1.15) | 0.61 | 1.13 (1.03, 1.24) | 0.010 | -0.29 (-0.36, -0.22) | <0.0001 | 0.08 (0.02, 0.14) | 0.0072 | -0.10 (-0.15, -0.04) | 0.00027 |
| Igbo | 0.31 (0.27, 0.35) | <0.0001 | 0.74 (0.62, 0.88) | 0.00059 | 0.37 (0.32, 0.43) | <0.0001 | 0.84 (0.75, 0.93) | <0.0001 | 0.24 (0.16, 0.32) | <0.0001 | 0.64 (0.57, 0.71) | <0.0001 |
| Ijaw | 0.41 (0.33, 0.50) | <0.0001 | 0.56 (0.41, 0.78) | 0.00054 | 0.42 (0.32, 0.54) | <0.0001 | 0.69 (0.54, 0.84) | <0.0001 | 0.45 (0.32, 0.57) | <0.0001 | 0.69 (0.57, 0.80) | <0.0001 |
| Kalenjin | 0.98 (0.52, 1.82) | 0.94 | 0.66 (0.32, 1.33) | 0.24 | 0.82 (0.50, 1.36) | 0.45 | -0.01 (-0.40, 0.38) | 0.96 | 0.26 (0.04, 0.47) | 0.019 | 0.16 (-0.12, 0.44) | 0.27 |
| Kamba | 0.97 (0.52, 1.82) | 0.93 | 0.33 (0.16, 0.69) | 0.0030 | 0.49 (0.30, 0.83) | 0.0071 | 0.01 (-0.39, 0.40) | 0.98 | 0.56 (0.34, 0.78) | <0.0001 | 0.38 (0.10, 0.66) | 0.0079 |
| Kasai | 1.64 (0.88, 3.05) | 0.12 | 0.85 (0.42, 1.71) | 0.65 | 1.11 (0.67, 1.83) | 0.68 | -0.26 (-0.65, 0.12) | 0.18 | 0.24 (0.02, 0.45) | 0.030 | -0.02 (-0.30, 0.26) | 0.88 |
| Kikuyu | 0.64 (0.34, 1.19) | 0.16 | 0.35 (0.17, 0.72) | 0.0042 | 0.34 (0.21, 0.57) | <0.0001 | 0.25 (-0.14, 0.63) | 0.21 | 0.55 (0.33, 0.76) | <0.0001 | 0.50 (0.22, 0.78) | 0.00044 |
| Kisii | 0.69 (0.36, 1.30) | 0.25 | 0.40 (0.19, 0.87) | 0.021 | 0.48 (0.28, 0.82) | 0.0068 | 0.22 (-0.17, 0.62) | 0.27 | 0.49 (0.27, 0.72) | <0.0001 | 0.47 (0.18, 0.76) | 0.0013 |
| Lomwe | 1.29 (0.80, 2.08) | 0.30 | 0.35 (0.19, 0.64) | 0.00075 | 0.43 (0.28, 0.65) | <0.0001 | -0.11 (-0.41, 0.19) | 0.49 | 0.66 (0.49, 0.84) | <0.0001 | 0.38 (0.16, 0.60) | 0.00065 |
| Luhya | 0.72 (0.39, 1.35) | 0.31 | 0.30 (0.14, 0.62) | 0.0011 | 0.37 (0.22, 0.62) | 0.00016 | 0.22 (-0.17, 0.61) | 0.26 | 0.60 (0.39, 0.81) | <0.0001 | 0.54 (0.26, 0.82) | 0.00014 |
| Luo | 0.67 (0.36, 1.25) | 0.21 | 0.30 (0.14, 0.61) | 0.0010 | 0.34 (0.21, 0.57) | <0.0001 | 0.37 (-0.02, 0.76) | 0.064 | 0.61 (0.40, 0.82) | <0.0001 | 0.64 (0.36, 0.92) | <0.0001 |
| Mandinka | 0.96 (0.88, 1.05) | 0.37 | 0.87 (0.76, 0.99) | 0.034 | 0.88 (0.80, 0.98) | 0.017 | 0.00 (-0.06, 0.07) | 0.90 | 0.10 (0.04, 0.15) | 0.00042 | 0.06 (0.02, 0.11) | 0.0070 |
| Mende | 1.01 (0.80, 1.27) | 0.96 | 1.16 (0.81, 1.68) | 0.42 | 0.96 (0.73, 1.26) | 0.77 | -0.12 (-0.28, 0.05) | 0.17 | 0.15 (0.02, 0.28) | 0.028 | 0.01 (-0.12, 0.13) | 0.91 |
| Mijikenda | 0.95 (0.51, 1.79) | 0.88 | 0.51 (0.25, 1.06) | 0.071 | 0.60 (0.36, 1.01) | 0.054 | 0.03 (-0.37, 0.43) | 0.88 | 0.35 (0.13, 0.57) | 0.0020 | 0.24 (-0.04, 0.53) | 0.095 |
| Mole-Dagbani | 0.57 (0.30, 1.08) | 0.085 | 0.74 (0.36, 1.52) | 0.42 | 0.57 (0.34, 0.95) | 0.033 | 0.34 (-0.06, 0.73) | 0.093 | 0.12 (-0.10, 0.34) | 0.28 | 0.25 (-0.03, 0.54) | 0.081 |
| Mossi | 0.92 (0.76, 1.10) | 0.35 | 0.82 (0.64, 1.04) | 0.10 | 0.80 (0.66, 0.98) | 0.030 | 0.07 (-0.08, 0.21) | 0.36 | 0.08 (-0.04, 0.20) | 0.18 | 0.10 (-0.01, 0.20) | 0.072 |
| Ngoni | 1.36 (0.85, 2.17) | 0.20 | 0.49 (0.28, 0.85) | 0.012 | 0.48 (0.32, 0.71) | 0.0003 | -0.16 (-0.46, 0.13) | 0.28 | 0.74 (0.57, 0.91) | <0.0001 | 0.40 (0.19, 0.61) | 0.00023 |
| Sara | 0.43 (0.29, 0.64) | <0.0001 | 0.42 (0.25, 0.69) | 0.00072 | 0.41 (0.27, 0.61) | <0.0001 | 0.44 (0.15, 0.72) | 0.0027 | 0.59 (0.38, 0.81) | <0.0001 | 0.73 (0.51, 0.94) | <0.0001 |
| Senoufo | 1.16 (1.00, 1.34) | 0.053 | 0.92 (0.75, 1.13) | 0.41 | 1.00 (0.85, 1.18) | 0.96 | -0.16 (-0.27, -0.05) | 0.0054 | 0.08 (-0.02, 0.17) | 0.10 | -0.04 (-0.12, 0.04) | 0.33 |
| Serere | 0.84 (0.75, 0.95) | 0.0054 | 0.67 (0.56, 0.80) | <0.0001 | 0.67 (0.59, 0.77) | <0.0001 | 0.08 (-0.00, 0.15) | 0.054 | 0.11 (0.04, 0.18) | 0.0011 | 0.11 (0.06, 0.17) | 0.00013 |
| Soninke/Serahuleh | 0.90 (0.79, 1.04) | 0.16 | 0.89 (0.74, 1.07) | 0.23 | 0.81 (0.69, 0.94) | 0.0059 | 0.00 (-0.10, 0.10) | 0.98 | 0.10 (0.01, 0.18) | 0.024 | 0.07 (-0.00, 0.14) | 0.063 |
| Temne | 0.92 (0.72, 1.17) | 0.50 | 1.07 (0.73, 1.56) | 0.72 | 0.99 (0.75, 1.31) | 0.93 | -0.02 (-0.19, 0.15) | 0.82 | 0.04 (-0.10, 0.18) | 0.57 | -0.00 (-0.13, 0.13) | 0.97 |
| Tiv | 0.46 (0.38, 0.56) | <0.0001 | 0.45 (0.34, 0.61) | <0.0001 | 0.31 (0.24, 0.39) | <0.0001 | 0.50 (0.35, 0.65) | <0.0001 | 0.61 (0.48, 0.74) | <0.0001 | 0.72 (0.60, 0.83) | <0.0001 |
| Tonga | 1.11 (0.70, 1.78) | 0.65 | 0.25 (0.14, 0.44) | <0.0001 | 0.44 (0.29, 0.65) | <0.0001 | -0.16 (-0.45, 0.14) | 0.30 | 0.63 (0.46, 0.80) | <0.0001 | 0.35 (0.13, 0.56) | 0.0013 |
| Tumbuka | 1.32 (0.82, 2.11) | 0.25 | 0.29 (0.16, 0.51) | <0.0001 | 0.44 (0.29, 0.65) | <0.0001 | -0.24 (-0.53, 0.06) | 0.12 | 0.78 (0.61, 0.95) | <0.0001 | 0.40 (0.19, 0.61) | 0.00025 |
| Ubangi and Itimbiri | 1.09 (0.58, 2.07) | 0.79 | 0.86 (0.41, 1.81) | 0.70 | 0.73 (0.43, 1.25) | 0.25 | 0.02 (-0.38, 0.43) | 0.91 | 0.30 (0.07, 0.54) | 0.011 | 0.21 (-0.08, 0.50) | 0.16 |
| Wollof | 0.87 (0.80, 0.95) | 0.0013 | 0.85 (0.76, 0.96) | 0.01 | 0.78 (0.71, 0.86) | <0.0001 | 0.06 (0.00, 0.12) | 0.039 | 0.07 (0.02, 0.12) | 0.0056 | 0.07 (0.03, 0.11) | 0.0010 |
| Xichangana | 0.72 (0.30, 1.71) | 0.46 | 0.19 (0.07, 0.51) | 0.0012 | 0.29 (0.15, 0.60) | 0.0007 | 0.12 (-0.42, 0.66) | 0.66 | 0.92 (0.63, 1.21) | <0.0001 | 0.68 (0.30, 1.06) | 0.00052 |
| Yoruba | 0.58 (0.53, 0.65) | <0.0001 | 0.65 (0.56, 0.75) | <0.0001 | 0.53 (0.47, 0.60) | <0.0001 | 0.37 (0.29, 0.45) | <0.0001 | 0.26 (0.20, 0.33) | <0.0001 | 0.37 (0.31, 0.43) | <0.0001 |
| ***Covariables*** |  |  |  |  |  |  |  |  |  |  |  |  |
| Accessibility to cities | 1.03 (0.99, 1.07) | 0.12 | 1.04 (0.99, 1.10) | 0.16 | 1.06 (1.02, 1.10) | 0.006 | 0.00 (-0.02, 0.03) | 0.79 | -0.03 (-0.05, -0.01) | 0.011 | -0.02 (-0.04, -0.00) | 0.030 |
| Age in days | 1.00 (1.00, 1.00) | <0.0001 | 1.00 (1.00, 1.00) | <0.0001 | 1.00 (1.00, 1.00) | <0.0001 | -0.00 (-0.00, -0.00) | <0.0001 | -0.00 (-0.00, -0.00) | <0.0001 | -0.00 (-0.00, -0.00) | <0.0001 |
| Caregiver secondary education | 0.87 (0.84, 0.89) | <0.0001 | 0.92 (0.87, 0.97) | 0.0024 | 0.88 (0.84, 0.91) | <0.0001 | 0.11 (0.09, 0.14) | <0.0001 | 0.03 (0.01, 0.05) | 0.0023 | 0.08 (0.07, 0.10) | <0.0001 |
| DPT-3 vaccination | 0.95 (0.92, 0.99) | 0.0060 | 0.98 (0.93, 1.04) | 0.56 | 0.96 (0.92, 1.00) | 0.078 | -0.00 (-0.03, 0.02) | 0.75 | -0.06 (-0.08, -0.04) | <0.0001 | -0.04 (-0.06, -0.02) | <0.0001 |
| Diarrhoea in past 2 weeks | 1.24 (1.20, 1.28) | <0.0001 | 1.27 (1.21, 1.34) | <0.0001 | 1.44 (1.39, 1.50) | <0.0001 | -0.21 (-0.23, -0.18) | <0.0001 | -0.19 (-0.21, -0.17) | <0.0001 | -0.24 (-0.26, -0.22) | <0.0001 |
| Finished floor material | 0.86 (0.83, 0.89) | <0.0001 | 1.00 (0.95, 1.05) | 0.92 | 0.90 (0.87, 0.94) | <0.0001 | 0.11 (0.08, 0.13) | <0.0001 | -0.01 (-0.03, 0.01) | 0.37 | 0.06 (0.04, 0.07) | <0.0001 |
| Household wealth index | 0.69 (0.67, 0.72) | <0.0001 | 0.76 (0.72, 0.81) | <0.0001 | 0.66 (0.64, 0.69) | <0.0001 | 0.26 (0.24, 0.28) | <0.0001 | 0.12 (0.10, 0.14) | <0.0001 | 0.23 (0.21, 0.25) | <0.0001 |
| ITN use night before | 0.94 (0.91, 0.96) | <0.0001 | 0.97 (0.93, 1.02) | 0.21 | 0.95 (0.92, 0.98) | 0.0036 | 0.03 (0.01, 0.05) | 0.0091 | 0.00 (-0.02, 0.02) | 0.85 | 0.02 (0.00, 0.03) | 0.02 |
| Improved sanitation | 0.92 (0.89, 0.95) | <0.0001 | 0.98 (0.93, 1.03) | 0.40 | 0.94 (0.91, 0.98) | 0.0018 | 0.07 (0.04, 0.09) | <0.0001 | 0.02 (0.00, 0.04) | 0.045 | 0.05 (0.03, 0.07) | <0.0001 |
| Improved water | 0.95 (0.93, 0.98) | 0.0023 | 1.00 (0.95, 1.04) | 0.87 | 0.97 (0.94, 1.00) | 0.085 | 0.02 (-0.00, 0.04) | 0.064 | -0.01 (-0.03, 0.01) | 0.41 | 0.01 (-0.01, 0.02) | 0.52 |
| LST ≥35⁰C | 0.95 (0.91, 1.00) | 0.034 | 1.12 (1.05, 1.20) | 0.00092 | 1.08 (1.03, 1.14) | 0.0041 | 0.01 (-0.02, 0.05) | 0.45 | -0.13 (-0.16, -0.10) | <0.0001 | -0.08 (-0.11, -0.05) | <0.0001 |
| Male | 1.30 (1.27, 1.33) | <0.0001 | 1.21 (1.17, 1.26) | <0.0001 | 1.24 (1.21, 1.28) | <0.0001 | -0.19 (-0.20, -0.17) | <0.0001 | -0.04 (-0.06, -0.03) | <0.0001 | -0.11 (-0.12, -0.10) | <0.0001 |
| Measles vaccination | 1.29 (1.25, 1.34) | <0.0001 | 1.01 (0.96, 1.07) | 0.68 | 1.07 (1.03, 1.12) | 0.00063 | -0.18 (-0.21, -0.16) | <0.0001 | -0.05 (-0.07, -0.03) | <0.0001 | -0.04 (-0.06, -0.03) | <0.0001 |
| Measurement position | 0.85 (0.82, 0.88) | <0.0001 | 0.69 (0.65, 0.73) | <0.0001 | - | - | 0.23 (0.21, 0.26) | <0.0001 | 0.23 (0.21, 0.25) | <0.0001 | - | - |
| Rainfall | 0.97 (0.90, 1.05) | 0.48 | 0.77 (0.68, 0.87) | <0.0001 | 0.81 (0.74, 0.89) | <0.0001 | 0.05 (0.00, 0.11) | 0.040 | 0.08 (0.03, 0.12) | 0.00059 | 0.08 (0.04, 0.12) | 0.00015 |
| Urban cluster | 0.86 (0.82, 0.89) | <0.0001 | 1.03 (0.97, 1.10) | 0.32 | 0.89 (0.85, 0.94) | <0.0001 | 0.15 (0.12, 0.18) | <0.0001 | 0.00 (-0.03, 0.03) | 0.98 | 0.09 (0.06, 0.11) | <0.0001 |
| Vegetation index | 1.06 (1.01, 1.11) | 0.016 | 0.84 (0.77, 0.90) | <0.0001 | 0.96 (0.91, 1.02) | 0.24 | -0.07 (-0.10, -0.03) | 0.00012 | 0.07 (0.04, 0.10) | <0.0001 | 0.01 (-0.02, 0.04) | 0.41 |

CI: Confidence intervals; HAZ: height-for-age z score; LST: land surface temperature OR: Odds Ratio; WAZ: weight-for-age z score; WHZ: weight-for-height z score.

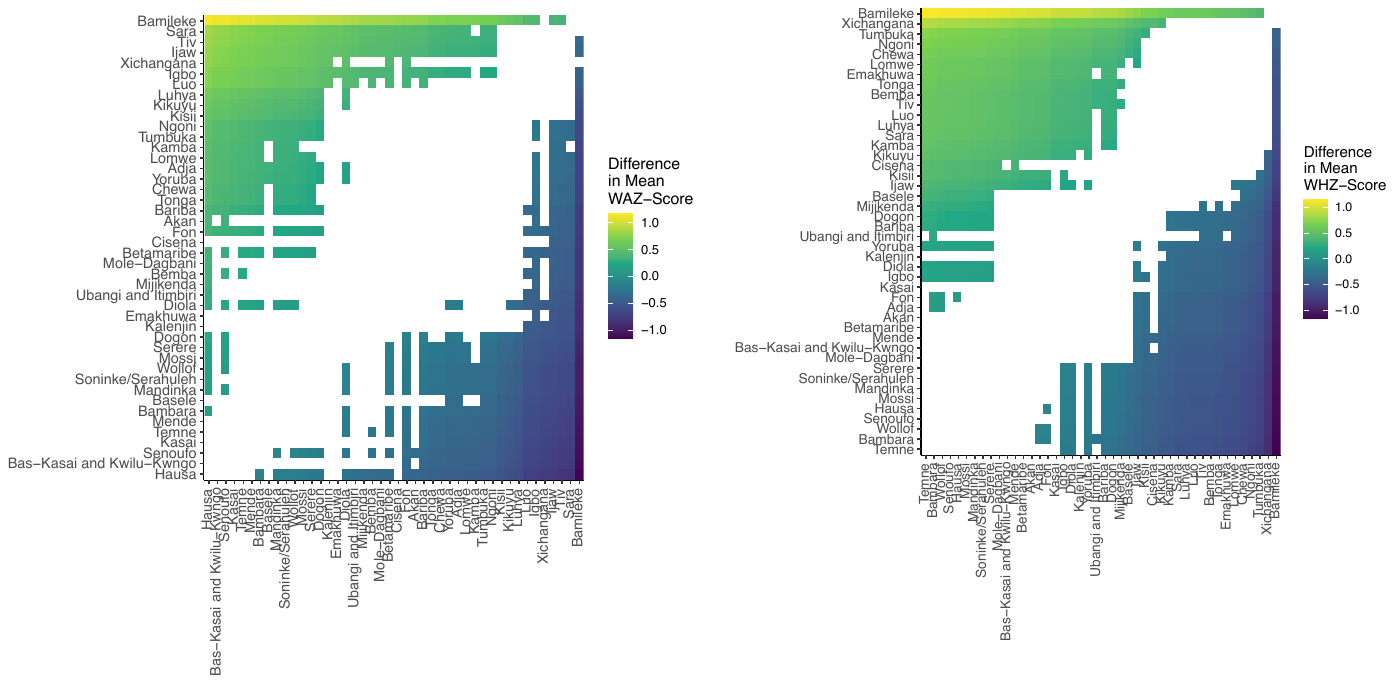

**Figure S1**. Growth variation relative to ethnic group among 138,312 children aged <5 years in 18 countries in sub-Saharan Africa surveyed between 2006 to 2019. Figures show significant pair-wise comparisons of mean weight-for-height Z scores and weight-for-age Z scores between ethnic groups.

**Figure S2.** Model validation for height-for-age (HAZ) z score. Calibration plots for the Gaussian and Binomial models. AUC for individual based predictions 0.74

**Figure S3.** Model validation for weight-for-height (WHZ) z score. Calibration plots for the Gaussian and Binomial models. AUC for individual based predictions 0.79

**Figure S4.** Model validation for weight-for-age (WAZ) z score. Calibration plots for the Gaussian and Binomial models. AUC for individual based predictions 0.75
